# Quantifying prior probabilities for disease-causing variants reveals the top genetic contributors in inborn errors of immunity

**DOI:** 10.1101/2025.03.25.25324607

**Authors:** Quant Group, Simon Boutry, Ali Saadat, Maarja Soomann, Johannes Trück, D. Sean Froese, Jacques Fellay, Sinisa Savic, Luregn J. Schlapbach, Dylan Lawless

## Abstract

**Background:** Accurate interpretation of genetic variants requires a quantitative estimate of how likely a variant is to contribute to disease, accounting for both observed and unobserved causal alleles across different inheritance modes.

**Methods:** We developed a statistical framework that computes genome-wide prior probabilities for variant classification by integrating population allele frequencies, disease classifications, and Hardy-Weinberg expectations across dominant, recessive, and X-linked inheritance. Bayesian modelling then combines these priors with individual-level data to produce credible intervals that quantify diagnostic confidence.

**Results:** The framework replaces categorical variant classification with continuous posterior probabilities that capture residual uncertainty from incomplete or missing genotype data. Demonstrations in three diagnostic scenarios show accurate quantification of variant-level disease relevance. Application to 557 genes implicated in inborn errors of immunity (IEI) generated a public database of prior probabilities. Integration with protein-protein interaction and immunophenotypic data revealed gene-level constraint patterns, and validation in national cohorts showed close agreement between predicted and observed case numbers.

**Conclusions:** Our method addresses a long-standing gap in clinical genomics by quantifying both observed and unobserved genetic evidence in disease diagnosis. It provides a reproducible probabilistic foundation for variant interpretation, clinical decision-making, and large-scale genomic analysis. ^1^

**Availability:** This data is integrated in public panels at https://iei-genetics.github.io. The source code are accessible as part of the variant risk estimation project at https://github.com/DylanLawless/var_risk_est and IEI-genetics project at https://iei-genetics.github.io. The data is available from the Zenodo repository: https://doi.org/10.5281/zenodo.15111583 (Var-RiskEst PanelAppRex ID 398 gene variants.tsv). VarRiskEst is available under the MIT licence.

**Graphical abstract:** 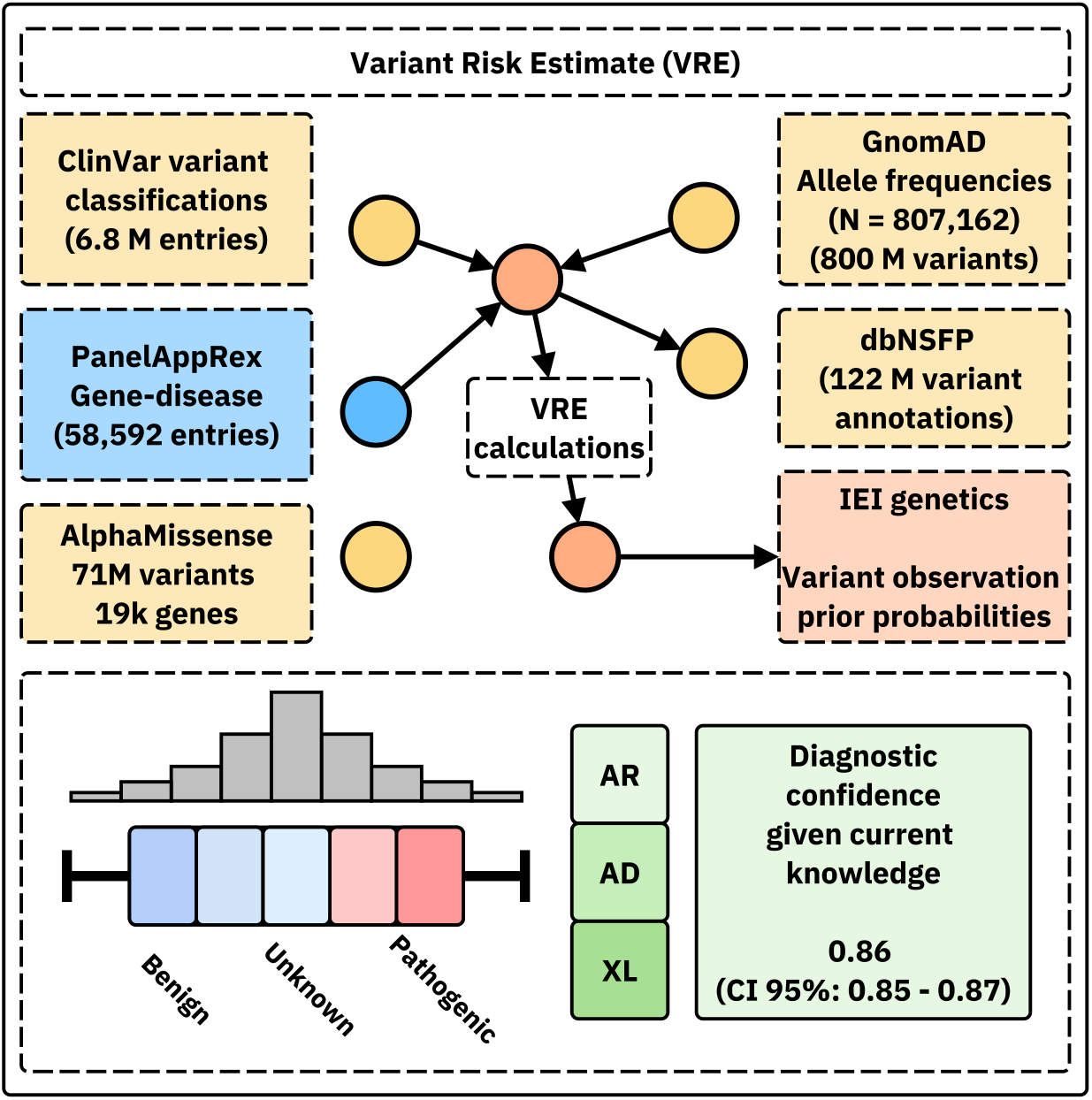

## 1 Introduction

Accurately estimating the probability that a patient carries a disease-causing genetic variant remains a central challenge in clinical and statistical genetics. For over a century, attention has focused on identifying variants which are true positive (TP) and reducing false positive (FP), while false negative (FN) and true negative (TN) have received comparatively little consideration. FN arise when pathogenic variants are missed due to technical or interpretive limits, and TN represent the vast background of benign variants. Although TN have clear value in contexts such as cancer screening, their wider statistical and clinical relevance remains underused. From a probabilistic standpoint, FN and TN provide crucial information about what is not observed, what should be expected under baseline genetic assumptions, and how confident one can be in the absence of a pathogenic finding. In practice, variant interpretation still relies on deterministic tools. For instance, GATK is used for processing; Ensembl VEP, SnpEff, FAVOR, and WGSA perform variant effect prediction (1); and Exomiser and VarFish support interpretation (2; 3). These tools focus on annotation and prioritisation but don’t have access to the underlying probabilities.

Quantifying the risk that a patient inherits a disease-causing variant is a core problem in genomics. Classical approaches based on Hardy-Weinberg Equilibrium (HWE) (4; 5) have long provided the foundation for calculating variant probabilities. However, these models become more complex when extended to different Mode of Inheritance (MOI), such as Autosomal Recessive (AR), Autosomal Dominant (AD), or X-Linked (XL) disorders. In AR diseases, risk depends on both homozygosity and compound heterozygosity, whereas a single allele can cause disease in AD or XL forms. Modern evidence shows that MOI can be further modulated by mechanisms including dominant negative effects, haploinsufficiency, mosaicism, and digenic or epistatic interactions (6). Although Karczewski et al. (7) advanced the field considerably, applying statistical genomics data across all MOI for any gene-disease combination remains unresolved. Recent disease-specific estimates, including Wilson disease, mucopolysaccharidoses, primary ciliary dyskinesia, and treatable metabolic disorders (8; 9), as reviewed by Hannah et al. (10), demonstrate progress but remain limited in scope.

All existing approaches remain limited to single MOI, specific disorders, or narrow gene sets. An integrated model is needed to unify these perspectives and generate probabilities that can serve as informative priors in a Bayesian framework for variant and disease probability estimation. Such a framework extends classical HWE-based methods, strengthens clinical interpretation by clarifying expected genetic outcomes, and enhances analytical models used in statistical genomics.

The resulting dataset from these necessary calculations also holds value for AI and reinforcement learning applications, providing an enriched version of the data underpinning frameworks such as AlphaFold (11) and AlphaMissense (12).

This gap has persisted largely due to the historical absence of harmonised reference datasets. Only recently have resources become available to enable rigorous genome-wide probability estimation, including population allele frequencies (gnomAD v4 (7)), curated variant classifications (ClinVar (13)), functional annotations (UniProt (14)), and pathogenicity prediction models (AlphaMissense (12)). We previously introduced PanelAppRex to integrate gene panel data from sources such as Genomics England (GE) PanelApp, ClinVar, and Universal Protein Resource (UniProt), enabling structured, machine-readable datasets for variant discovery and interpretation (13–16). Together, these advances now allow systematic modelling of the expected distribution of variant types, frequencies, and classifications across the genome.

In this study, we report genome-wide probabilities of disease observation across gene-disease combinations, focusing on known Inborn Errors of Immunity (IEI) genes, also referred to as Primary Immunodeficiency (PID) or monogenic inflammatory bowel disease genes (15–17). This well-defined genotype-phenotype set, comprising more than 500 gene-disease associations, provides a robust validation of clinical relevance. The latest International Union of Immunological Societies (IUIS) classification and diagnostic guidelines (17; 18) serve as the benchmark reference. We hypothesised that by integrating curated annotations on population Allele Frequency (AF), disease phenotypes, MOI patterns, and variant classifications with HWE-based calculations, it would be possible to estimate expected probabilities of disease-associated variants and derive quantitative confidence in genetic diagnosis using these priors.

## 2 Results

### 2.1 Occurrence probability across disease genes

Our study integrated large-scale annotation databases with gene panels from PanelAppRex to systematically assess disease genes by MOI (15). By combining population allele frequencies with ClinVar clinical classifications, we computed an expected occurrence probability for each SNV, representing the likelihood of encountering a variant of a specific pathogenicity for a given phenotype. We report these probabilities for 54,814 ClinVar variant classifications across 557 genes (linked dataset (19)).

We focused on panels related to Primary Immunodeficiency or Monogenic Inflammatory Bowel Disease, using PanelAppRex panel ID 398. **Figure 1** displays all reported ClinVar variant classifications for this panel. The resulting natural scaling system (-5 to +5) accounts for the frequently encountered combinations of classification labels (e.g. benign to pathogenic). The resulting dataset (19) is briefly shown in **Table 1** to illustrate that our method yielded estimates of the probability of observing a variant for every particular ClinVar classification.

**Table 1:** Example output from the framework estimating variant occurrence probability for disease-associated genes. The table shows the first several rows of results for 557 genes from PanelAppRex’s panel (ID 398: Primary immunodeficiency or monogenic inflammatory bowel disease). “ClinVar Significance” lists the pathogenicity classification assigned by ClinVar, while “Occurrence Prob” represents the calculated probability of observing each variant class given the phenotype. The mode of inheritance (MOI) is gene-disease specific. Additional data, such as population allele frequency, are omitted for brevity. (19)

| Gene | Panel ID | ClinVar Clinical Sig | GRCh38 Pos | HGVSc | HGVSp | MOI | Occurrence Prob |
| --- | --- | --- | --- | --- | --- | --- | --- |
| ABI3 | 398 | Uncertain significance | 3:49210771 | c.47G>A | p.Arg16Gln | AR | 0.000000007 |
| ABI3 | 398 | Uncertain significance | 3:49216678 | c.265C>T | p.Arg89Cys | AR | 0.000000005 |
| ABI3 | 398 | Uncertain significance | 3:49217742 | c.289G>A | p.Val97Met | AR | 0.000000002 |
| ABI3 | 398 | Uncertain significance | 3:49217781 | c.328G>A | p.Gly110Ser | AR | 0.000000002 |
| ABI3 | 398 | Uncertain significance | 3:49217844 | c.391C>T | p.Pro131Ser | AR | 0.000000015 |
| ABI3 | 398 | Uncertain significance | 3:49220257 | c.733C>G | p.Pro245Ala | AR | 0.000000022 |
| ... | ... | ... | ... | ... | ... | ... | ... |

**Figure 1:**
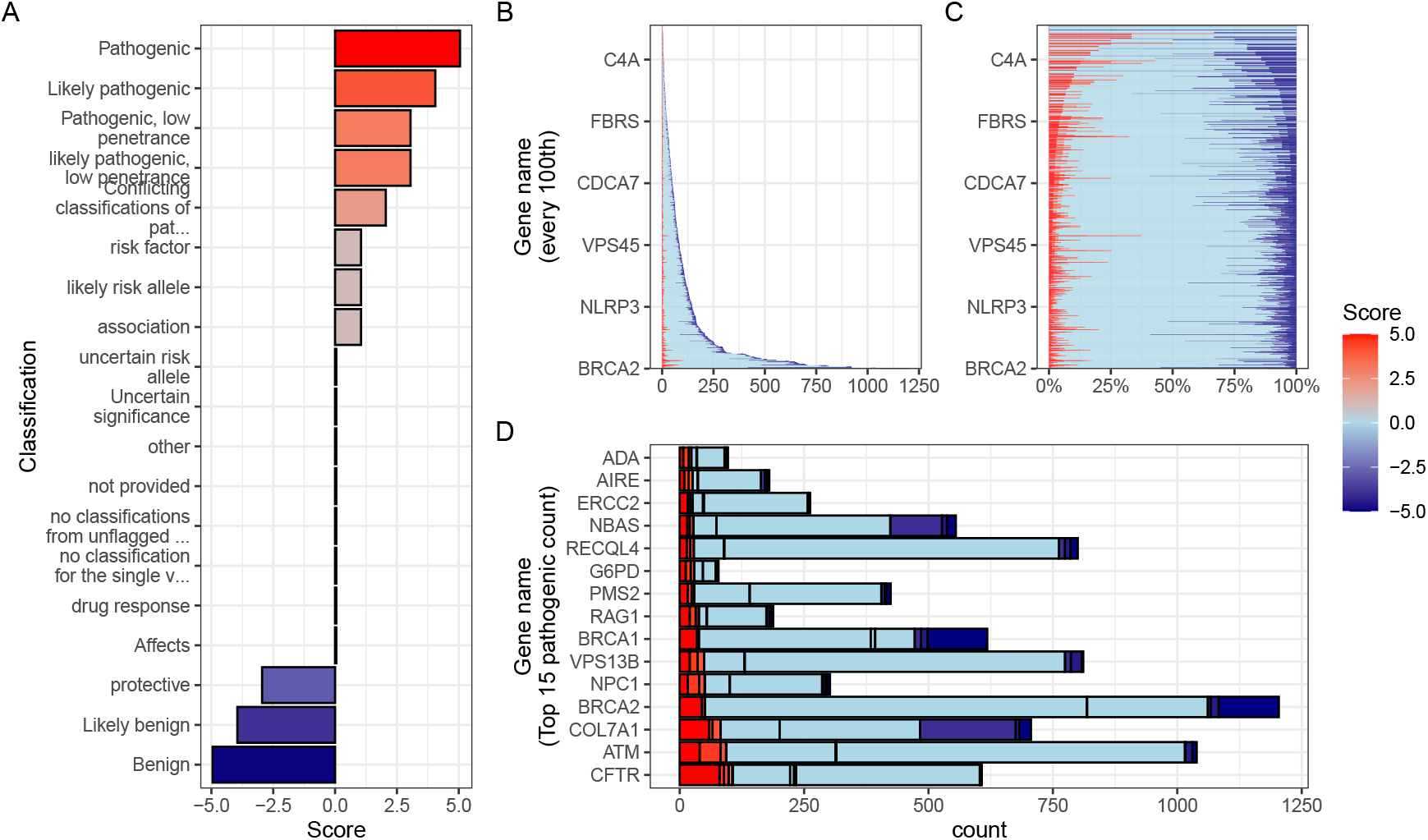
Summary of ClinVar clinical significance classifications in the PID gene panel. (A) Shows the numeric score coding for each classification (i.e. -5 benign to +5 pathogenic) as defined in methods Section 5.2. (B) Displays the stacked absolute count of classifications per gene. The same counts are shows as percentages per gene in (C). (D) For demonstration, the top 15 genes ranked by absolute count of pathogenic (score 5) variant classifications, indicating those most frequently occurring in the population as disease-causing.

### 2.2 Integrating observed true positives and unobserved false negatives into a single, actionable conclusion

We applied the probabilistic framework to an example patient to illustrate its diagnostic utility. The model assessed all known variant positions, identifying reference sites as TN, observed variants as TP, and unsequenced or low-quality sites as potential FN. By modelling both observed and unobserved evidence, the framework evaluated each variant in relation to all others to derive an evidence-weighted probability that a damaging allele is present in the genome. While the framework quantifies all variant classifications, the examples here focus on causal pathogenicity.

### 2.3 Scenario one - complete coverage and simple diagnosis

We present the results from three scenarios for an example single-case patient being investigated for the genetic diagnosis of IEI with severe autoinflammatory disease. **Figure S2** shows the results of the first simple scenario, in which only one known pathogenic variant, Nuclear Factor Kappa B Subunit 1 (*NFKB1*) `p.Ser237Ter`, was observed and all other previously reported genome-wide pathogenic positions were successfully sequenced and confirmed as reference. In this setting, the model assigned the full posterior probability to the observed allele, yielding 100 % confidence that all present evidence supported a single, true positive causal explanation. The most strongly supported observed variant was `p.Ser237Ter` (posterior: 0.594). The strongest (probability of observing) non-sequenced variant was a benign variant `p.Thr567Ile` (posterior: 0). The total probability of a causal diagnosis given the available evidence was 1 (95% CI: 1–1) (**Table S1**).

### 2.4 Scenario two - incomplete coverage and complex diagnosis

**Figure 2** illustrates the second scenario, where the same pathogenic *NFKB1* variant `p.Ser237Ter` was detected, but sequencing coverage was incomplete at three additional classified sites. Among these was the likely pathogenic splice-site variant `c.159+1G>A` , which was not captured in the sequencing data. The panels in **Figure 2 (A-F)** show the incremental integration of observed and missing evidence, resulting in a posterior probability that reflects both confirmed findings and residual uncertainty. Supporting metrics and reporting conclusions are summarised in **Table S2** and **Table 2**.

**Table 2:**
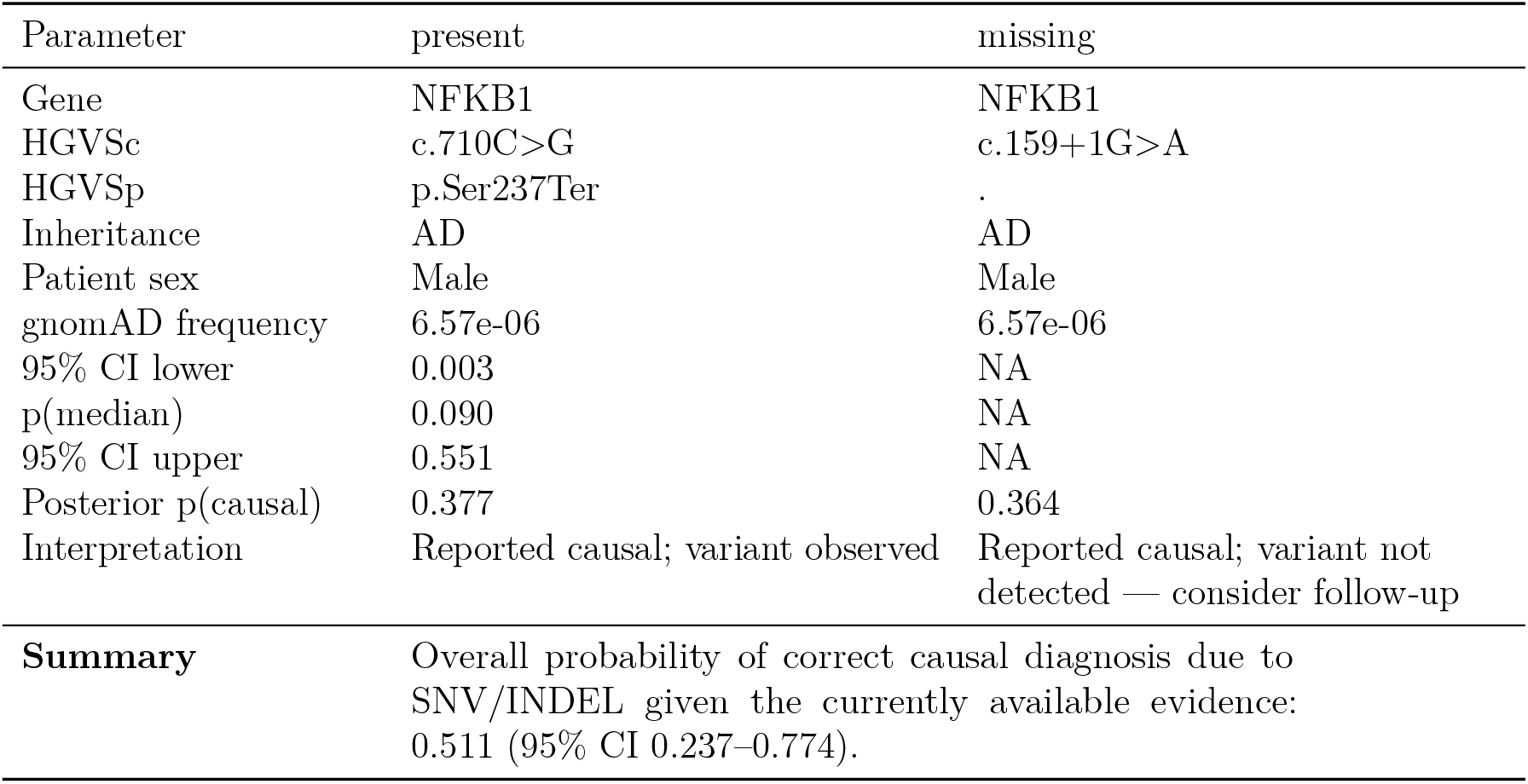
Final variant report for clinical genetics scenario 2. Reported causal: p.Ser237Ter (posterior 0.377). Undetected causal: c.159+1G>A (posterior 0.364). The total probability of a causal diagnosis given the available evidence was 0.511 (95% CI: 0.237–0.774).

| Parameter | present | missing |
| --- | --- | --- |
| Gene | NFKB1 | NFKB1 |
| HGVSc | c.710C>G | c.159+1G>A |
| HGVSp | p.Ser237Ter | . |
| Inheritance | AD | AD |
| Patient sex | Male | Male |
| gnomAD frequency | 6.57e-06 | 6.57e-06 |
| 95% CI lower | 0.003 | NA |
| p(median) | 0.090 | NA |
| 95% CI upper | 0.551 | NA |
| Posterior p(causal) | 0.377 | 0.364 |
| Interpretation | Reported causal; variant observed | Reported causal; variant not detected — consider follow-up |
| **Summary** | Overall probability of correct causal diagnosis due to SNV/INDEL given the currently available evidence: 0.511 (95% CI 0.237–0.774). |  |

**Figure 2:**
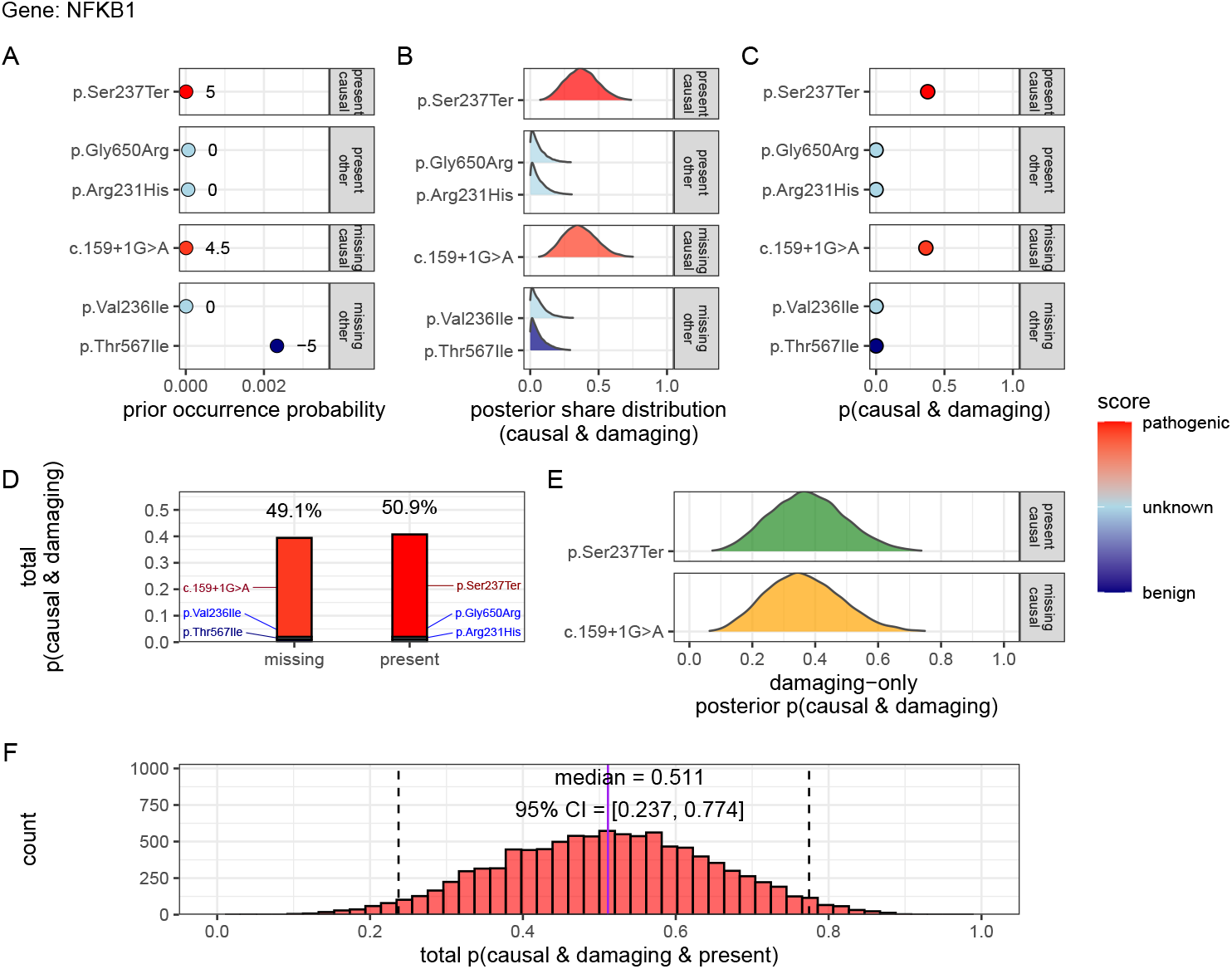
Quantifying diagnostic certainty from genomic evidence. Example of *NFKB1* (scenario 2) with observed (TP) and missing (FN) causal variants. The proband carried three heterozygous variants, including pathogenic `p.Ser237Ter` , with incomplete coverage at three loci, including splice-site variant `c.159+1G>A`. Panels show: (A) prior occurrence probabilities; (B) posterior variant weights 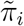; (C) pervariant posterior probability of being damaging and causal; (D) posterior for causal variants; (E) decomposition of total pathogenic probability into observed (green) and missing (orange) components; and (F) the final posterior probability that a damaging allele is present (median 0.54, 95 % CrI 0.26-0.80). Results are compared to **Table 2**.

Bayesian integration of all annotated alleles produced quantitative Credible Interval (CrI)s for pathogenic attribution that (i) preserve Hardy-Weinberg expectations, (ii) accommodate AD, AR, and XL inheritance, and (iii) quantify uncertainty from unsequenced or failed-Quality Control (QC) sites. **Figure 2 (A)** depicts the prior distribution partitioned by observed or missing status and by causal evidence, coloured by ClinVar score. **Figure 2 (B)** shows posterior normalisation, concentrating probability density on two high-evidence alleles. **Figure 2 (C)** displays the per-variant posterior probability of being damaging and causal, highlighting the confirmed nonsense variant `p.Ser237Ter` and the missing splice-donor `c.159+1G>A`. **Figure 2 (D)** restricts the view to causal candidates, showing posterior mass across these two variants. **Figure 2 (E)** decomposes total damaging probability into observed (40,%) and missing (34,%) components, and **Figure 2 (F)** summarises the gene-level posterior, with a median probability of 0.542 (95,% CrI: 0.264-0.8).

Numerically, the present variant `p.Ser237Ter` contributes 0.399 of the posterior share and the missing splice-donor allele `c.159+1G>A` contributes 0.339, with remaining alleles contributing negligibly (**Table S2**). For this patient scenario, the probability of correct genetic diagnosis due to *NFKB1* `p.Ser237Ter` is 0.542 (95,% CrI: 0.264-0.8), reflecting the remaining uncertainty from an unconfirmed alternative allele. If the second variant is confirmed absent, the confidence increases to 1 (95,% CrI: 1-1), consistent with scenario one.

### 2.5 Scenario three - currently impossible diagnosis

**Figure S3** shows the third scenario, where no observed variants were detected in the proband for *NFKB1*. Instead, a broad range of plausible FNs were identified as missing for Tumor necrosis factor, alpha-induced protein 3 (*TNFAIP3*). The strongest of these unsequenced variants was `p.Cys243Arg` (posterior: 0.347). However, the total probability of a causal diagnosis for the patient *given the available evidence* was 0 (95,% CI: 0-0), since these missing variants must be explicitly accounted for (**Table S3**). Upon confirmation, these probabilities can update as shown in scenario two.

### 2.6 Posterior probabilities quantify competing variant hypotheses

The three scenarios deliberately remove the distraction of other genome-wide candidate variants, which are assumed as present and accounted for within the model. In practice, the same posterior probability and CrI calculations are performed across all qualifying variants. In real-world diagnostics, we commonly find multiple variants to carry non-negligible probabilities. Our framework explicitly quantifies these competing hypotheses, enabling a ranked interpretation that reflects the totality of evidence. Overlapping CrI do not indicate ambiguity in the method, but rather a principled measure of remaining uncertainty. This output can directly inform follow-up actions, such as functional validation or treatment trials, and supports the use of CrI thresholds as a transparent decision-making aid when data are incomplete or equivocal.

### 2.7 Quantifying variant probabilities improves classification of inborn errors of immunity

#### Genes with high pathogenic variant burden show structured network constraint

We applied the framework to IEI, a disease area which offers a well-curated gene set to validate genome-wide estimates and demonstrate potential applications (17). Because pathogenicity in IEI often reflects shared molecular pathways, we integrated ClinVar-derived variant probabilities with Protein-Protein Interaction (PPI) data to quantify per-gene pathogenic burden and assess whether constraint clusters within specific networks, consistent with known IEI categories and immunophenotypes (7; 20).

ClinVar classifications (**Figure 1**) were scaled from -5 to +5 by pathogenicity. We summed all positive (potentially damaging) scores per gene to derive the score-positive-total metric. **Figure S9 (A)** shows the PPI network, with node size and colour proportional to this metric (log-transformed). The top 15 genes with the highest total prior probabilities of disease association are labelled, consistent with **Figure 1**.

One-way Analysis of Variance (ANOVA) indicated that major IEI categories differed in score-positive-total values (*F* (8, 500) = 2.82, *p* = 0.0046), suggesting uneven distributions of pathogenic variant burden (**Figure S9 (B)**). However, Tukey Honestly Significant Difference (HSD) post hoc tests (**Figure S9 (C)**) showed overlapping 95% Confidence Interval (CI)s for all pairwise comparisons, indicating that category-level differences were not individually significant.

#### Constrained subnetworks reveal distinct functional disease signatures

To visualise the PPI network, we applied Uniform Manifold Approximation and Projection (UMAP) projection (**Figure 3**). Node size indicates connectivity, and colours show cluster membership (20). Blue labels mark hub genes (top 5% by degree), and yellow labels mark the 15 genes with the highest damaging variant load. Genes enriched for pathogenic variants were distinct from highly connected nodes, indicating that Loss-of-Function (LOF) in hubs is constrained, while damaging variants in less connected genes have more specific effects.

**Figure 3:**
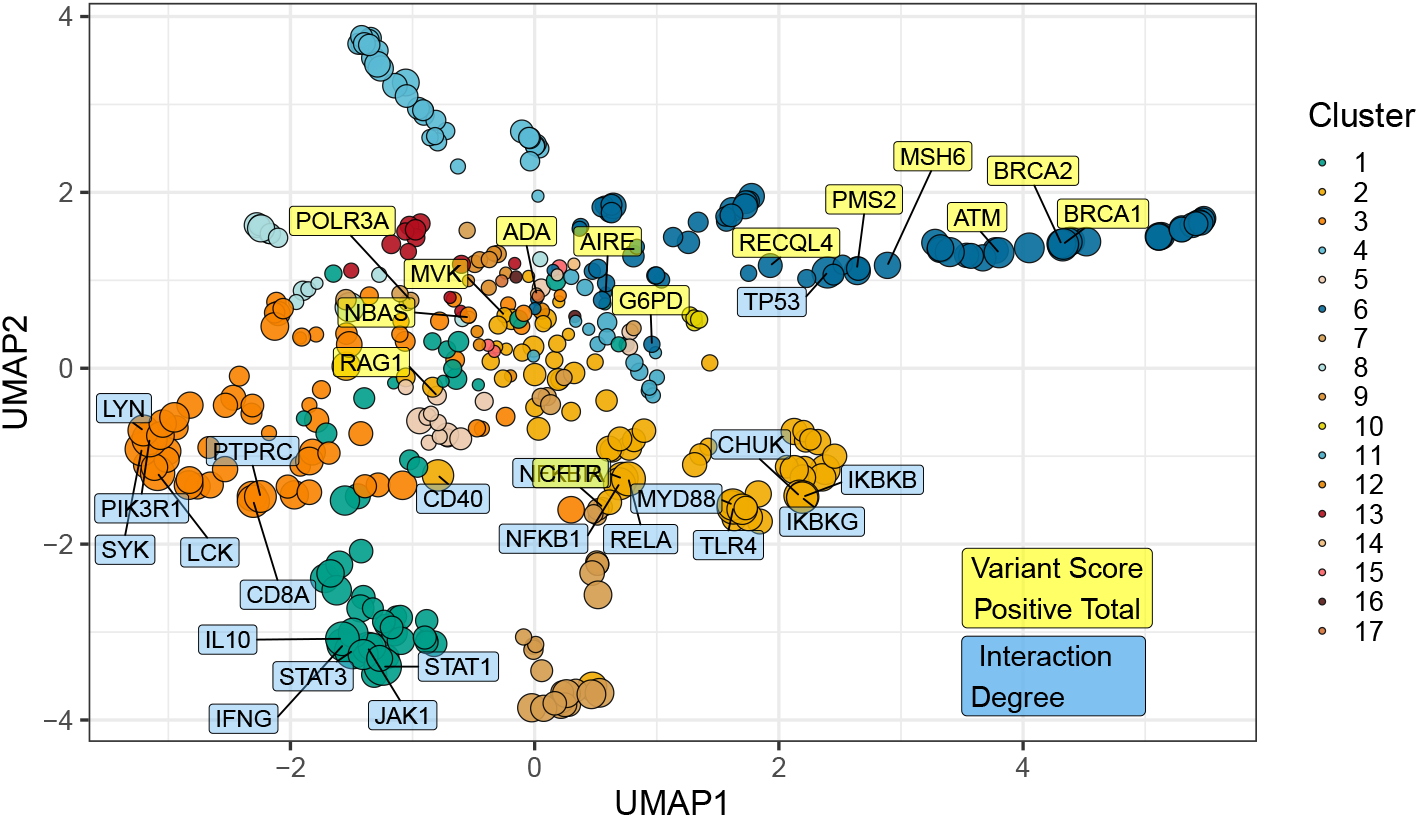
UMAP embedding of the PPI network. Projection of the high-dimensional protein interaction network into two dimensions. Nodes are coloured by cluster and scaled by interaction degree. Blue labels denote hub genes (degree above the 95th percentile) and yellow labels mark the top 15 genes by score-positive-total (damaging ClinVar classifications). The separation indicates that genes with high pathogenic variant loads differ from highly connected nodes.

**Figure S10** shows standardised residuals for major disease categories across clusters corresponding to **Figure 3**. The dendrogram groups categories with similar enrichment patterns, and the bar plot highlights the strongest signals. Complement Deficiencies (CD) (8) and Bone Marrow Failure (BMF) (9) showed the highest enrichment, indicating that their genes cluster in subnetworks with a high burden of damaging variants. All categories exceeded nominal significance (|residual| *>* 2), reflecting concentrated variant effects in specific clusters, notably cluster 4 for category 8.

Building on these findings, we tested the relationship between PPI connectivity (degree) and LOF constraint (Loss-Of-function Observed/Expected Upper bound Fraction (LOEUF) rank) (7). A weak but significant negative correlation (*ρ* = −0.181, *p* = 0.00024) indicated that highly connected genes tend to be more constrained. Cluster-level analyses revealed stronger relationships, for example cluster 2 (*ρ* = −0.375, *p* = 0.000994) and cluster 4 (*ρ* = −0.800, *p <* 0.000001), suggesting shared mechanisms of LOF intolerance within specific pathways (**Figure S12**). The score-positive-total metric effectively summarised aggregate pathogenic burden and selective constraint.

Functional enrichment using MsigDB and FUMA (21) linked clusters 2 and 4 to distinct disease pathways (**Figure S13**). Cluster 2 was enriched for inflammatory and autoimmune traits, including ankylosing spondylitis, psoriasis, inflammatory bowel disease, and rheumatoid arthritis. Cluster 4 was associated with complement and ocular phenotypes, such as macular degeneration, complement biomarkers, nephropathy, and pulmonary traits. GTEx v8 tissue expression data (**Figure S14**) supported these patterns, showing that variant burden and network constraint together define mechanistic disease relationships that extend established classifications.

#### Accounting for genome-wide relationships and linkage disequilibrium

**Figure 4 (A)** presents a genome-wide karyoplot of all IEI panel genes mapped to GRCh38, coloured by MOI. Panels **(B)** and **(C)** show regional locus plots for *NFKB1* and *CFTR*. In **(B)**, only benign variants such as p.Ala475Gly display high observation probabilities in the AD gene *NFKB1*, which is highly intolerant to LOF. In contrast, **(C)** shows elevated probabilities for pathogenic variants in *CFTR*, consistent with its well-established disease association. Analysis of Linkage Disequilibrium (LD) using R^2^ demonstrates that regions of high linkage can be effectively modelled, enabling separation of independent variant signals within complex loci.

**Figure 4:**
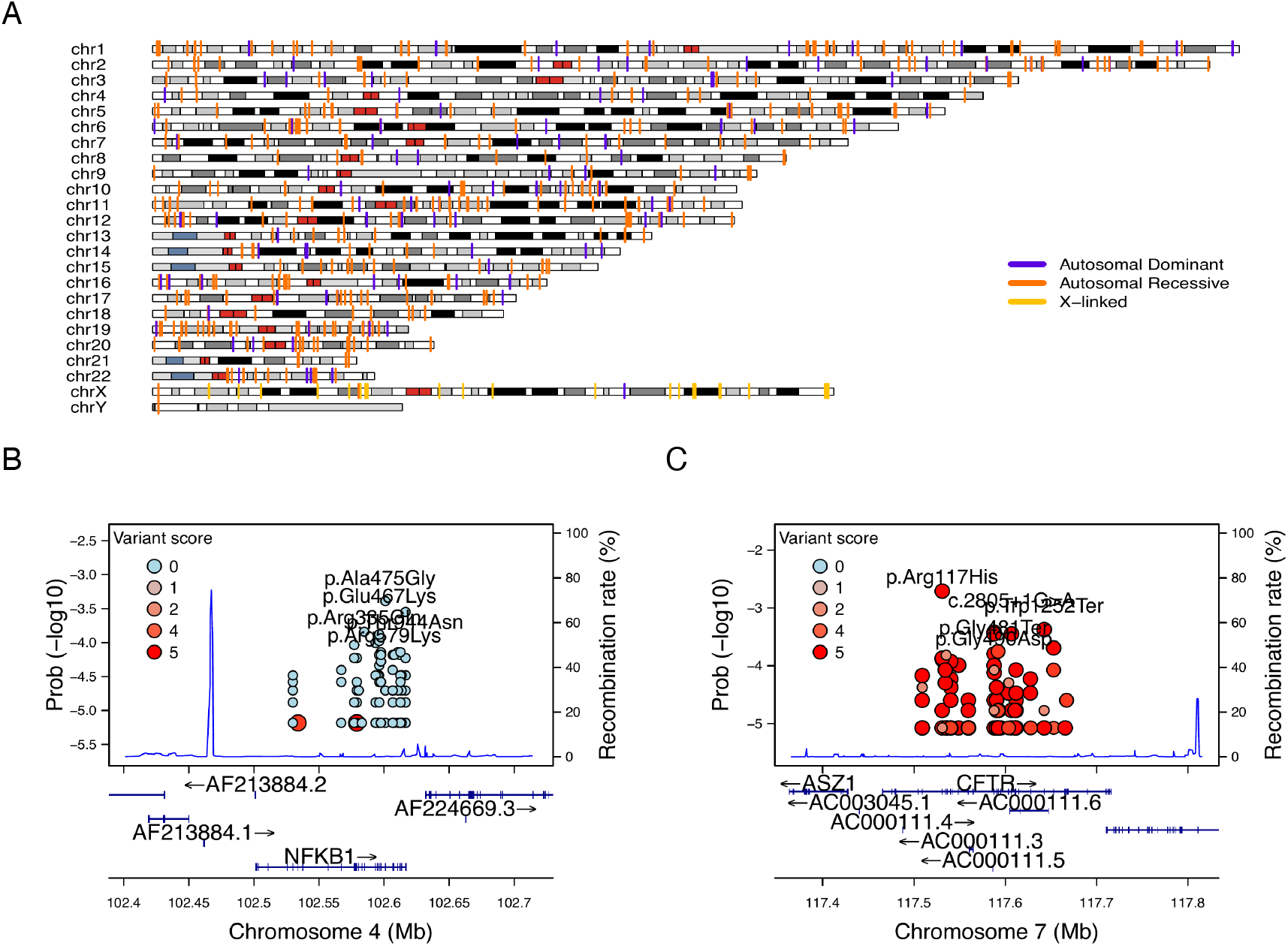
Genome-wide IEI, variant occurrence probability and LD by R^2^. (A) Genome-wide karyoplot of all IEI panel genes mapped to GRCh38, with colours indicating MOI. (B) Zoomed-in locus plot example for *NFKB1* showing variant occurrence probabilities; only benign variants such exhibit high probabilities in this AD gene intolerant to LOF. (C) Locus plot example for *CFTR* displaying high probabilities for pathogenic variants; due to the dense clustering of pathogenic variants, score filter >0 was applied. Top five variants are labelled per gene.

#### Data-driven classification of PID genes

Immunophenotypic data from the IUIS IEI tables link genes to clinical immune phenotypes but are variable and incomplete. They nonetheless provide a foundation for reproducible, data-driven classification of IEI genes. Using the estimated probabilities of disease observation from variant classification likelihoods, we simplified 315 immunophenotypic features from free-text descriptions into binary categories (normal vs. not-normal) for T cells, B cells, Immunoglobulin (Ig), and neutrophils (**Figure S15**). Mapping these features to STRINGdb PPI clusters showed significant non-random associations (*χ*^2^ *<* 1*e*−13; **Figure S16**), indicating that network structure captures major immunophenotypic patterns. Predictive accuracy rose from 33% using phenotypes alone to 61% when combined with IUIS categories (**Figure S17**), with Ig and T-cell abnormalities as top predictors. The final PPI-immunophenotype model, dependent on the underlying probability of disease observation, defined 17 data-driven PID groups (33 with full IUIS categories; **Figure 5**). This approach advances classification from qualitative expert assessment to reproducible, data-driven grouping of related disorders.

**Figure 5:**
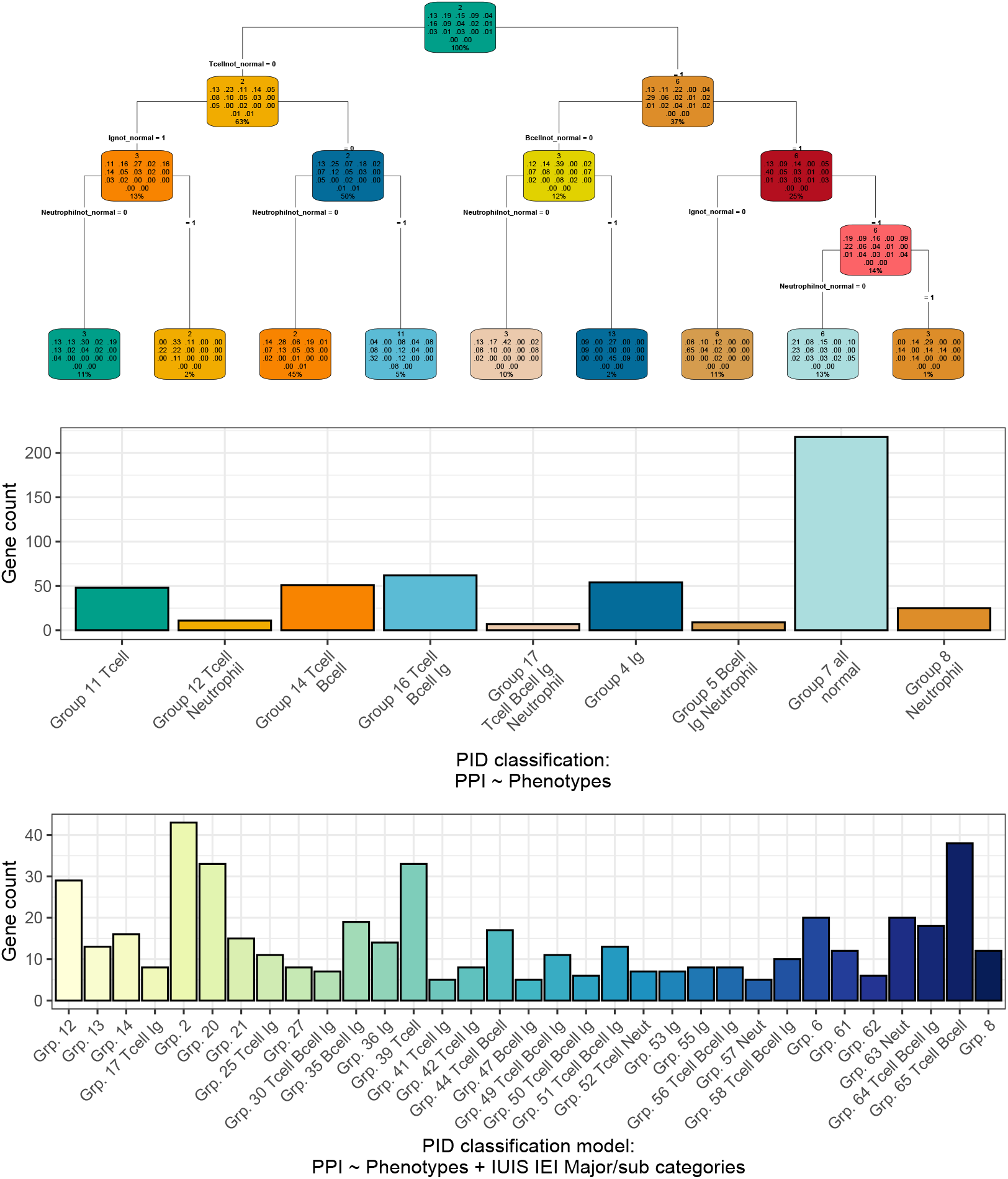
Data-driven model integrating variant probability and refining PID classifications. (Top) Decision tree grouping genes into 17 new PID classes based on immunophenotypic, PPI, and variant occurrence features, including LOEUF constraint. Each node summarises gene count, class probabilities, and sample fraction. (Middle) Distribution of the 17 PID classes, labelled by predominant abnormal immune features (e.g. T cell, B cell, Ig, Neutrophil). (Bottom) Extended model incorporating traditional IUIS IEI categories. This quantitative taxonomy unites variant probability, gene constraint, and immune phenotype into a scalable, machine-readable framework.

### 2.8 Observed variant likelihood is independent of AlphaMissense pathogenicity

AlphaMissense (and similar tools) assigns pathogenicity scores to all possible amino acid substitutions, yet **Figure 6** shows that variants most frequently observed in patients are predominantly benign or of uncertain significance. This highlights the gap between predicting pathogenic potential and estimating how likely a variant is to occur, providing a quantitative basis for improving predictive models. Comparison of **Figure 6 (A)** with **Figure 1 (D)** illustrates the distribution of classifications. A Kruskal-Wallis test (**Figure S18**) found no meaningful differences in variant observation probability across AlphaMissense classification groups. This indicates that current pathogenicity predictors estimate how damaging a variant may be, but not how likely it is to be observed in patients. The weak inverse trends likely reflect underlying effects of MOI and LOF intolerance.

**Figure 6:**
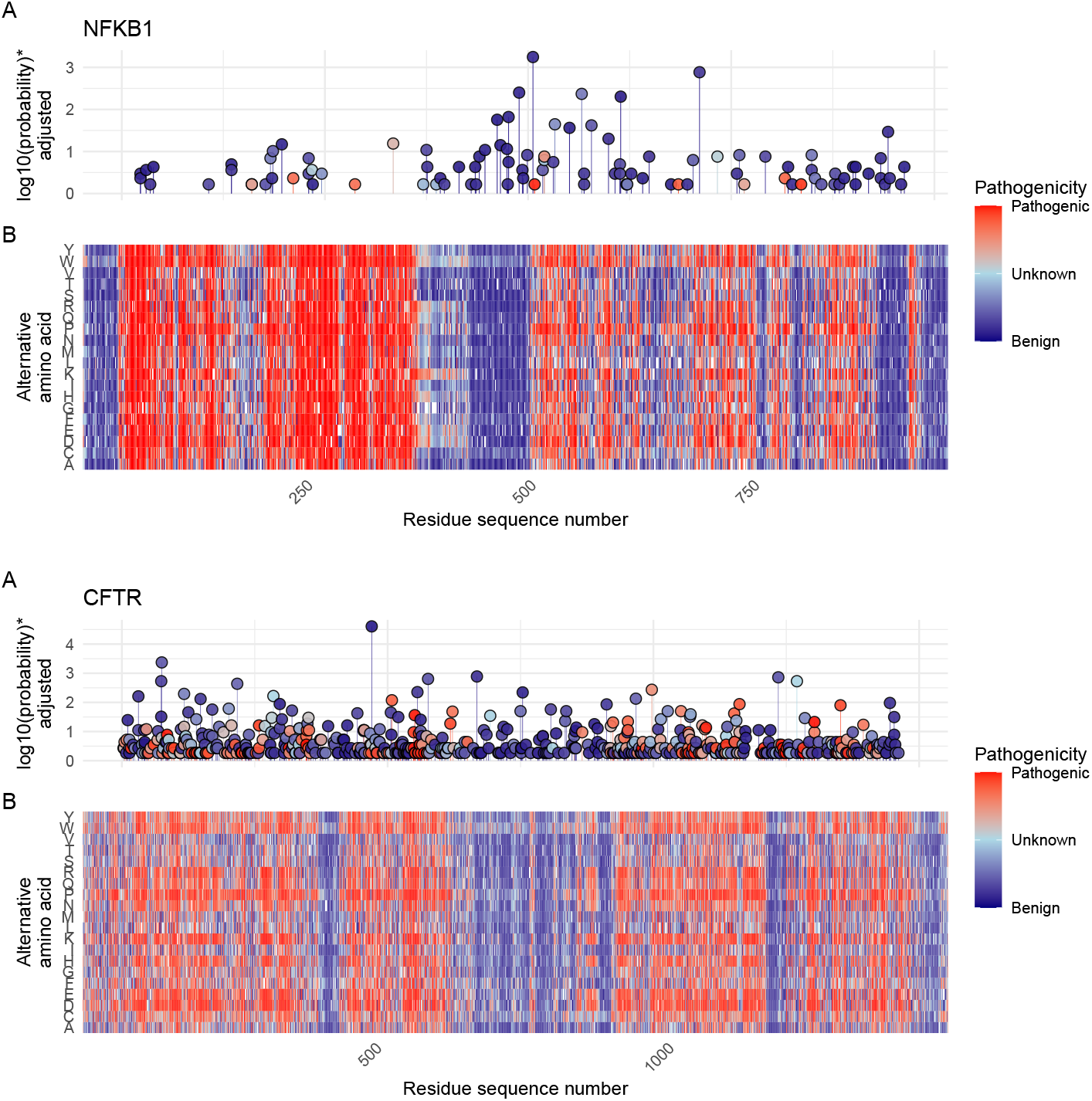
Variant observation probabilities and AlphaMissense pathogenicity scores. For each gene, the upper plot (A) displays variant observation probabilities and the lower plot (B) shows AlphaMissense-derived pathogenicity scores across all amino acid substitutions. Although AlphaMissense predicts the functional impact of every substitution, the variants most frequently observed in patients are mainly benign or of uncertain significance. This illustrates that pathogenicity prediction captures functional effect but not likelihood of occurrence, highlighting the complementary nature of these probability measures. *Axis scaling adjusted for visibility near zero; higher values indicate higher observation probability.

### 2.9 Integration of variant observation probabilities into the IEI genetics framework

We integrated prior probabilities of variant observation across all IEI genes linked to each phenotype (17), spanning AD, AR, and XL MOI. These gene-level priors, derived from curated PanelAppRex panels, quantify disease-gene relationships and extend the IEI genetics framework. The outputs include machine- and human-readable datasets and a web interface (https://iei-genetics.github.io) integrating these probabilities with clinical and functional data (**Figure S19**). Each record displays summary statistics, sparkline box plots, and dynamic links to external resources (e.g. ClinVar, Online Mendelian Inheritance in Man (OMIM), AlphaFold) for direct bioinformatic use (19).

## 3 Discussion

We present, to our knowledge, the first genome-wide framework for quantifying prior probabilities of observing disease-associated variants and for performing evidence-aware genetic diagnosis with CrI (8; 10). By integrating population allele frequencies (Genome Aggregation Database (gnomAD)) (7), curated classifications (ClinVar) (13), and functional annotations (database for Non-Synonymous Functional Predictions (dbNSFP)), with classical HWE-based models, we derived probabilistic estimates for 54,814 ClinVar variant classifications across 557 IEI genes linked to PID and monogenic inflammatory bowel disease (15; 17). Although demonstrated in IEI, the framework generalises to all inheritance modes: AD and XL requiring a single pathogenic allele, and AR involving homozygous or compound heterozygous variants. These HWE-based priors provide a baseline for Bayesian inference, now feasible at scale through comprehensive genomic databases (7; 12; 13; 15).

A persistent limitation in clinical genetics is the focus on confirming only observed TP variants. Our framework addresses this by integrating both observed and unobserved evidence into a single quantitative conclusion. We derived per-variant priors across all MOI and combined these with the patient’s genotype data to yield posterior probabilities representing the likelihood of carrying a damaging causal allele. This approach provides a clinically interpretable probability that unifies confirmed and potentially missed variants. Incidentally, the score-positive-total metric offers a practical, evidence-based method for ranking candidate genes and guiding diagnosis in phenotypically overlapping cases.

Our current framework had several limitations. It focuses primarily on Single Nucleotide Variant (SNV)s and does not yet incorporate structural variants, de novo mutations, hypomorphic alleles, overdominance, variable penetrance, tissue-specific effects, or phenomena such as the Wahlund effect and pleiotropy (6). Future extensions could include factors such as embryonic lethality, condition-specific penetrance, and age of onset (10). The present analysis also assumes random mating, large population size, and no migration, mutation, or selection. We demonstrated genome-wide gene distribution and MOI for the IEI panel in relation to LD, confirming both feasibility and relevance. However, accurate LD estimation requires population-specific, genomewide pairwise genotype data. Our use of global reference allele frequencies improves generalisability but may reduce precision relative to population-specific estimates. The integration of IUIS-derived immunophenotypic data in the novel PID groupings assumes that simplified categorisation improves their suitability for computational rather than intuitive interpretation. Although the underlying IUIS annotations are variable and incomplete, and in some disorders such as GATA2 deficiency reflect secondary rather than primary effects, this approach represents a step toward data-driven rather than intuition-based disease classification.

Across diagnostic scenarios, the framework quantified the probability that any variant, regardless of classification, is both observed and relevant to disease. This enabled confident attribution to present variants while accounting for counterfactual and unsequenced but plausible sites. Scenario two exemplified how confirmed and missing evidence jointly shape diagnostic confidence. Unlike conventional approaches that consider only observed findings, the framework captures residual uncertainty and produces structured, “evidence-aware” conclusions. Combined with genome-wide priors from PanelAppRex, it provides a scalable foundation for probabilistic, evidence-aware diagnostics across phenotypes.

Estimating genetic disease risk is complicated by uncertainty in penetrance and the fraction of cases attributable to specific variants (6). In probabilistic terms, *P* (*D*) is the population risk of disease, *P* (*G*) is the frequency of a given genotype, *P* (*D* | *G*) is the penetrance (the probability of disease given that genotype), and *P* (*G* | *D*) is the proportion of affected individuals carrying that genotype. In the simplest case of a single, fully penetrant allele, the lifetime risk *P* (*D*) equals the genotype frequency *P* (*G*), given by *P* (*D*) = *p*^2^ for autosomal recessive and *P* (*D*) = 2*p*(1 − *p*) ≈ 2*p* for autosomal dominant inheritance. With incomplete penetrance *P* (*D* | *G*), the relationship becomes

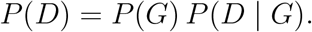

When multiple variants contribute, the proportion of cases attributable to each variant is described by *P* (*G* | *D*), giving

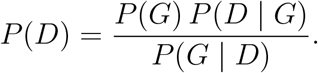

Both *P* (*D* | *G*) and *P* (*G* | *D*) are rarely known precisely (22; 23). Our framework addresses this by integrating variant classifications, allele frequencies, and curated gene-disease associations. While individual estimates are uncertain, their aggregate behaviour is well-calibrated under James-Stein estimation principles (24), allowing reliable priors for Bayesian inference.

Curated gene-disease associations identify loci that explain most cases, permitting approximation of *P* (*G* | *D*) ≈ 1 and stable estimation of *P* (*G*), the frequency of disease-associated genotypes. For a filtered subset of variants 𝒱, where each has probability *P* (*G*_*i*_ | *D*), we assume ∑ _*i*∈ 𝒱_ *P* (*G*_*i*_ | *D*) ≈ 1. Even if this sum is slightly less than one, resulting risk estimates remain robust within typical credible intervals. These pre-computed priors provide a quantitative foundation for refining disease probability and diagnostic confidence through Bayesian integration.

By aggregating variant sets beyond single-gene analyses, the framework complements established rare variant association tests such as Sequence Kernel Association Test (SKAT) and Aggregated Cauchy Association Test (ACAT) (25–28) and aligns with multi-omics integration methods (29; 30). It is compatible with American College of Medical Genetics and Genomics (ACMG) guidelines (31), related interpretation frameworks (32; 33), and current QC standards (34; 35). Standardised systems, including ACMG Secondary Findings v3.2 (36), allow integration of these probabilistic results into clinical workflows. Comparison with AlphaMissense scores showed that pathogenicity predictors estimate molecular effect rather than probability of occurrence. Integrating variant-level priors with predictors like AlphaMissense could improve disease relevance by incorporating gene-disease associations and MOI for applications in AI/ML.

Future work should extend this framework to additional variant classes and models, iteratively updating classical estimates with new data to refine probabilistic diagnostics and support more precise clinical decision-making.

## 4 Conclusion

We present a statistical framework that quantifies the probability that any variant contributes to disease by integrating observed genotypes with the possibility of unobserved alleles. It replaces binary classifications with continuous posterior probabilities and credible intervals reflecting evidence and uncertainty. Although illustrated in IEI, the approach is generalisable to all diseases and supports quantitative genomic interpretation at scale.

## Acknowledgements

We would like to thank all the patients and families who have been providing advice on SwissPedHealth and its projects, as well as the clinical and research teams at the participating institutions. We acknowledge Genomics England for providing public access to the PanelApp data. The use of data from Genomics England panelapp was licensed under the Apache License 2.0. The use of data from UniProt was licensed under Creative Commons Attribution 4.0 International (CC BY 4.0). ClinVar asks its users who distribute or copy data to provide attribution to them as a data source in publications and websites (13). dbNSFP version 4.4a is licensed under the Creative Commons Attribution-NonCommercial-NoDerivatives 4.0 International (CC BY-NC-ND 4.0); while we cite this dataset as used our research publication, it is not used for the final version which instead used ClinVar and gnomAD directly. GnomAD is licensed under Creative Commons Zero Public Domain Dedication (CC0 1.0 Universal). GnomAD request that usages cites the gnomAD flagship paper (7) and any online resources that include the data set provide a link to the browser, and note that tool includes data from the gnomAD v4.1 release. AlphaMissense asks to cite Cheng et al. (12) for usage in research, with data available from Cheng et al. (37).

## Contributions

DL designed the analyses and wrote the manuscript. SB, AS, MS, and JT designed analysis and wrote the manuscript. DSF, and SS wrote the manuscript. JF, LJS supervised the work, and applied for funding. The Quant Group is a collaboration across multiple institutions where authors contribute equally; the members on this project were DL, SB, and AS.

## Ethics statement

This study only used data which was previously published and publicly available, as cited in the manuscript. This SwissPedHealth study, under which this work was carried out, was approved based on the advice of the ethical committee Northwest and Central Switzerland (EKNZ, AO_2022-00018). The study was conducted in accordance with the Declaration of Helsinki.

## Data availability

The data used in this manuscript is derived from open sources which are cited in methods. The data generated is available from the Zenodo repository:. The resulting data are also reproducible using the code repository https://github.com/DylanLawless/var_risk_est.

## 5 Methods

### 5.1 Dataset

Data from gnomAD v4 comprised 807,162 individuals, including 730,947 exomes and 76,215 genomes (7). This dataset provided 786,500,648 SNVs and 122,583,462 Insertion/Deletion (InDel)s, with variant type counts of 9,643,254 synonymous, 16,412,219 missense, 726,924 nonsense, 1,186,588 frameshift and 542,514 canonical splice site variants. ClinVar data were obtained from the variant summary dataset (as of: 16 March 2025) available from the NCBI FTP site, and included 6,845,091 entries, which were processed into 91,319 gene classification groups and a total of 38,983 gene classifications; for example, the gene *A1BG* contained four variants classified as likely benign and 102 total entries (13). For our analysis phase we also used dbNSFP which consisted of a number of annotations for 121,832,908 SNVs (38). The PanelAppRex core model contained 58,592 entries consisting of 52 sets of annotations, including the gene name, disease-gene panel ID, diseases-related features, confidence measurements. PPI network data was provided by Search Tool for the Retrieval of Interacting Genes/Proteins (STRINGdb), consisting of 19,566 proteins and 505,968 interactions (20). The Human Genome Variation Society (HGVS) nomenclature is used with Variant Effect Predictor (VEP)-based codes for variant IDs. AlphaMissense includes pathogenicity prediction classifications for 71 million variants in 19 thousand human genes (12; 37). We used these scores to compared against the probability of observing the same given variants. **Box 1** list the definitions from the IUIS IEI for the major disease categories used throughout this study (17).

The following genes were used for disease cohort validations and examples. We used the two most commonly reported genes from the IEI panel *NFKB1* (39–42) and Cystic Fibrosis Transmembrane Conductance Regulator (*CFTR*) (43–45) to demonstrate applications in AD and AR disease genes, respectively. We used Severe Combined Immunodeficiency (SCID)-specific genes AR DNA Cross-Link Repair 1C (*DCLRE1C*), AR Recombination activating gene 1 (*RAG1*), XL Interleukin 2 Receptor Subunit Gamma (*IL2RG*) to demonstrate a IEI subset disease phenotype of SCID. We also used AD *TNFAIP3* for other examples comparable to *NFKB1* since it is also causes AD pro-inflammatory disease but has more known ClinVar classifications at higher AF then *NFKB1*.

#### Box 1: definitions

**Major Category Description**

1. CID Immunodeficiencies affecting cellular and humoral immunity
2. CID+ Combined immunodeficiencies with associated or syndromic features
3. PAD - Predominantly Antibody Deficiencies
4. PIRD - Diseases of Immune Dysregulation
5. PD - Congenital defects of phagocyte number or function
6. IID - Defects in intrinsic and innate immunity
7. AID - Autoinflammatory Disorders
8. CD - Complement Deficiencies
9. BMF - Bone marrow failure

### 5.2 Variant classification occurrence probability

To quantify the likelihood that an individual harbours a variant with a given disease classification, we compute the variant-level occurrence probability (variant risk estimate (VRE)) for each variant. As a starting point, we considered the classical HWE for a biallelic locus: *p*^2^ + 2*pq* + *q*^2^ = 1, where *p* is the allele frequency, *q* = 1 − *p, p*^2^ represents the homozygous dominant, 2*pq* the heterozygous, and *q*^2^ the homozygous recessive genotype frequencies. For disease phenotypes, particularly under AR MOI, the risk is traditionally linked to the homozygous state (*p*^2^); however, to account for compound heterozygosity across multiple variants, we allocated the overall gene-level risk proportionally among variants.

Our computational pipeline estimated the probability of observing a disease-associated genotype for each variant and aggregated these probabilities by gene and ClinVar classification. This approach included all variant classifications, not limited solely to those deemed “pathogenic”, and explicitly conditioned the classification on the given phenotype, recognising that a variant could only be considered pathogenic relative to a defined clinical context. The core calculations proceeded as follows:

#### 1. Allele frequency and total variant frequency

For each variant *i* in a gene, the allele frequency was denoted as *p*_*i*_. For each gene (any genomic region or set), we defined the total variant frequency (summing across all reported variants in that gene) as: *P*_tot_ =∑_*i*∈gene_ *p*_*i*_. Note that, because each calculation is confined to one gene, no additional scaling was required for our primary analyses (*P*_tot_). However, if this same unscaled summation is applied across regions or variant sets of differing size or dosage sensitivity, it can bias burden estimates. In such cases, normalisation by region length or incorporation of gene- or region-specific dosage constraints is recommended.

If any of the possible SNV had no observed allele (*p*_*i*_ = 0), we assigned a minimal risk: 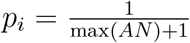 where max(*AN*) was the maximum allele number observed for that gene. This adjustment ensured that a nonzero risk was incorporated even in the absence of observed variants in the reference database.

#### 2. Occurrence probability based on MOI

The probability that an individual is affected by a variant depends on the MOI. For **AD** and **XL** variants, a single pathogenic allele suffices: *p*_disease,*i*_ = *p*_*i*_. For **AR** variants, disease manifests when two pathogenic alleles are present, either as homozygotes or as compound heterozygotes. We use: *p*_disease,*i*_= *p*_*i*_ *P*_tot_.

Under HWE, the overall gene-level probability of an AR genotype is 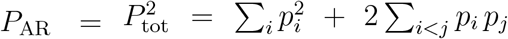 , where *P*_tot_ = ∑_*i*_ *p*_*i*_. A naïve per-variant assignment 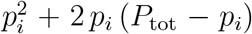 would, when summed over all *i*, double-count the compound heterozygous terms. To partition *P*_AR_ among variants without double counting, we allocate risk in proportion to each variant’s allele frequency: 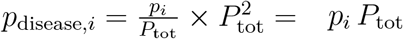.

This ensures 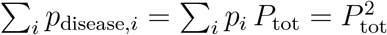 , recovering the correct AR risk while attributing each variant its fair share of homozygous and compound-heterozygous contributions.

More simply, for AD or XL conditions a single pathogenic allele suffices, so the classification risk (e.g. benign, pathogenic) equals its population frequency. For AR conditions two pathogenic alleles are required - either two copies of the same variant or one copy each of two different variants, so we divide the overall recessive risk among variants according to each variant’s share of the total classification frequency in that gene.

#### 3. Expected case numbers and case detection probability

Given a population with *N* births (e.g. as seen in our validation studies, *N* = 69 433 632), the expected number of cases attributable to variant *i* was calculated as: *E*_*i*_ = *N* · *p*_disease,*i*_. The probability of detecting at least one affected individual for that variant was computed as: *P* (≥ 1)_*i*_ = 1 – (1 – *p* _disease,*i*_)^*N*^.

#### 4. Aggregation by gene and ClinVar classification

For each gene and for each ClinVar classification (e.g. “Pathogenic”, “Likely pathogenic”, “Uncertain significance”, etc.), we aggregated the results across all variants. The classification grouping can be substituted by any alternative score system. The total expected cases for a given group was: *E*_group_ = ∑_*i*∈group_ *E*_*i*_, and the overall probability of observing at least one case within the group was calculated as: *P*_group_ = 1 − Π_*i*∈group_ (1 − *p*_disease,*i*_).

#### 5. Data processing and implementation

We implemented the calculations within a High-Performance Computing (HPC) pipeline and provided an example for a single dominant disease gene, *TNFAIP3*, in the source code to enhance reproducibility. Variant data were imported in chunks from the annotation database for all chromosomes (1-22, X, Y, M).

For each data chunk, the relevant fields were gene name, position, allele number, allele frequency, ClinVar classification, and HGVS annotations. Missing classifications (denoted by “.”) were replaced with zeros and allele frequencies were converted to numeric values. Subsequently, the variant data were merged with gene panel data from PanelAppRex to obtain the disease-related MOI mode for each gene. For each gene, if no variant was observed for a given ClinVar classification (i.e. *p*_*i*_ = 0), a minimal risk was assigned as described above. Finally, we computed the occurrence probability, expected cases, and the probability of observing at least one case of disease using the equations presented.

The final results were aggregated by gene and ClinVar classification and used to generate summary statistics that reviewed the predicted disease observation probabilities. We define the *VRE* as the prior probability of observing a variant classified as the cause of disease

#### 6. Score-positive-total

For use as a simple summary statistic on the resulting user-interface, we defined the *score-positive-total* as the total number of positively scored variant classifications within a given region (gene, locus, or variant set). Using the ClinVar classification assigned to a scale from−5 (benign) to +5 (pathogenic), we included only scores *>* 0, corresponding to some evidence of pathogenicity. The score-positive-total yields a non-normalised estimate of the prior probability that a phenotype is explained by known pathogenic variants.

#### 7. Classification scoring system

Each ClinVar classification was assigned an integer score: pathogenic = +5, likely pathogenic = +4, pathogenic (low penetrance) = +3, likely pathogenic (low penetrance) = +2, conflicting pathogenicity = +2, likely risk allele/risk factor/association = +1, drug response/uncertain significance/no classification/affects/other/not provided/uncertain risk allele = 0, protective = - 3, likely benign = -4, benign = -5. No further normalisation was applied. The resulting distribution (**Figure S1 A-B**) is naturally comparable to a zero-centred average rank (**C-D**). This straightforward, modular approach can be readily replaced by any comparable evidence-based classification system. Variants with scores ≤ 0 were omitted, since benign classifications do not inform disease likelihood in the score-positive-total summary.

### 5.3 Bayesian framework for posterior probability of genetic diagnosis

We developed a Bayesian framework to estimate the probability that at least one variant in a given genome or target set is both present and clinically relevant in a proband. The method combines variant-specific priors derived from any classification system or scoring scheme with a probabilistic model of genotype presence, incorporating both observed and unsequenced genomic positions.

#### Prior specification

Each variant *i* was assigned a prior probability *p*_*i*_ reflecting its likelihood of being relevant to the user’s target interpretation. This could include, for example, being classified as pathogenic, benign, or uncertain significance, or being prioritised by a quantitative scoring scheme. The priors may be derived from allele frequency, expert curation, functional prediction, or any other source of variant-level evidence. The framework supports modular priors without constraint. In our implementation, priors are derived from ClinVar classification, gnomAD population frequency, and MOI specified with PanelAppRex gene-disease curation. We modelled the latent causality of each variant as a Beta distribution parameterised by *α*_*i*_ = round(*p*_*i*_ · *A*_*N*_) + *w, β*_*i*_ = *A*_*N*_ − round(*p*_*i*_ · *A*_*N*_) + 1, where *A*_*N*_ is the total number of callable alleles at the site (e.g. *A*_*N*_ = 2*n* for *n* diploid individuals), and *w* is a small stabilising constant (typically *w* = 1). This construction reflects the expected frequency of the variant in the tested cohort under the assumption of causal relevance.

#### Posterior simulation

We drew *N* = 10,000 samples from the Beta prior to approximate a posterior distribution for each variant: 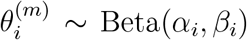 , *m* = 1, … , *N*. In each simulation round, these values were normalised across all variants to reflect the relative share of the total causal signal:

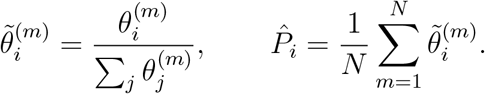

This yields a posterior expectation of variant-level contribution, which can be thresholded or ranked depending on the application.

#### Genotype presence model

Independently from the classification or interpretive context, each variant was assigned a probability *q*_*i*_ ∈ [0, 1] of being present in the proband. For observed alternate genotypes, *q*_*i*_ = 1; for confidently called reference genotypes, *q*_*i*_ = 0; and for unsequenced or uncertain sites, *q*_*i*_ was set to the prior probability *p*_*i*_, or an alternative estimate reflecting presence likelihood. For each simulation round, we drew a genotype indicator: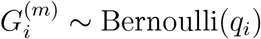 , and computed the total interpreted variant probability: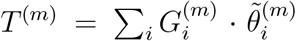. The empirical distribution of {*T* ^(*m*)^} represents the posterior belief that the proband harbours at least one variant matching the specified interpretive criteria in the given gene or set. Summary statistics, such as the posterior median and 95% credible interval, are used to report this probability with interval-based accountability.

#### Flexibility and implementation

This framework generalises across different scoring systems, prior models, and sequencing contexts. It can incorporate uncertain classifications, variant weights, unsequenced positions, or alternative genotype priors without altering the core inference structure. The method is implemented in R with modular inputs and reproducible simulation logic.

### 5.4 Application to diagnostic scenarios with observed and unobserved variant data

In this section, we detail our approach to integrating sequencing data with prior classification evidence (e.g. pathogenic on ClinVar) to calculate the posterior probability of a complete successful genetic diagnosis. Our method is designed to account for possible outcomes of TP, TN, and FN, by first ensuring that all nucleotides corresponding to known variant classifications (benign, pathogenic, etc.) have been accurately sequenced. This implies the use of genomic variant call format (gVCF)-style data which refer to variant call format (VCF)s that contain a record for every position in the genome (or interval of interest) regardless of whether a variant was detected at that site or not. Only after confirming that these positions match the reference alleles (or novel unaccounted variants are classified) do we calculate the probability that additional, alternative pathogenic variants (those not observed in the sequencing data) could be present. Our CrI for pathogenicity thus incorporates uncertainty from the entire process, including the tally of TP, TN, and FN outcomes. We ignore the contribution of FPs as a separate task to be tackled in the future.

We estimated, for every query (e.g. gene or disease-panel), the posterior probability that at least one constituent allele is both damaging and causal in the proband. The workflow comprises four consecutive stages.

#### (i) Data pre-processing

We synthesized an example patient in a disease cohort of 200 cases. We made several scenarios where a causal genetic diagnosis based on the available data is either simple, difficult, or impossible. Our example focused on a proband two representative genes for AD IEI: *NFKB1* and *TNFAIP3*. All coding and canonical splice-region variants for *NFKB1* were extracted from the gVCF. We assumed a typical QC scenario, where sites corresponding to previously reported pathogenic alleles were checked for read depth ≥ 10 and genotype quality ≥ 20. Positions that failed this check were labelled *missing*, thus separating true reference calls from non-sequenced or uninformative sequence.

#### (ii) Evidence mapping and occurrence probability

PanelAppRex variants were annotated with ClinVar clinical significance. Each label was converted to an ordinal evidence score *S*_*i*_ ∈ [−5, 5] and rescaled to a pathogenic weight *W*_*i*_ = rescale(*S*_*i*_; −5, 5 → 0, 1). This scoring system can be replaced with any comparable alternative. The HWE-based pipeline of Section 5.2 supplied a per-variant occurrence probability *p*_*i*_. The adjusted prior was 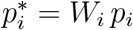 , and flag_*i*_ ∈ {present, missing}.

#### (iii) Prior specification

In a hypothetical cohort of *n* = 200 diploid individuals the count of allele *i* follows a Beta-Binomial model. Marginalising the Binomial yields the Beta prior 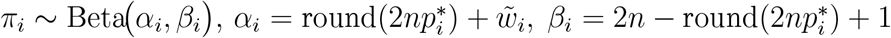 , where 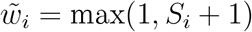 contributes an additional pseudo-count whenever *S*_*i*_ *>* 0.

#### (iv) Posterior simulation and aggregation

For each variant *i* we drew *M* = 10 000 realisations 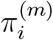 and normalised within each iteration, 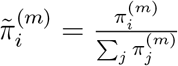.Variants with *S*_*i*_ *>* 4 were deemed to have evidence as *causal* (pathogenic or likely pathogenic). We note that an alternative evidence score or conditional threshold can be substituted for this step. Their mean posterior share 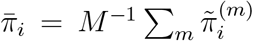 and 95% CrI were retained. The probability that a damaging causal allele is physically present was obtained by a second layer: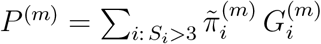 , and 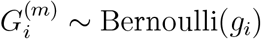 , with *g*_*i*_ = 1 for present variants, *g*_*i*_ = 0 for reference calls, and *g*_*i*_ = *p*_*i*_ for missing variants. The gene-level estimate is the median of 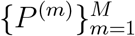 and its 2.5^th^/97.5^th^ percentiles.

#### (v) Scenario analysis

The three scenarios were explored for a causal genetic diagnosis that is either simple, difficult, or impossible given the existing data. The proband spiked data had either: (1) known classified variants only, including only one known TP pathogenic variant, *NFKB1* `p.Ser237Ter` , (2) inclusion of an additional plausible yet non-sequenced splice-donor allele *NFKB1* `c.159+1G>A` (likely pathogenic) as a FN, and (3) where no known causal variants were present for a patient, one representative variant from each distinct ClinVar classification was selected and marked as unsequenced to emulate a range of putative FNs. The selected variants were: *TNFAIP3* `p.Cys243Arg` (pathogenic), `p.Tyr246Ter` (likely pathogenic), `p.His646Pro` (conflicting interpretations of pathogenicity), `p.Thr635Ile` (uncertain significance), `p.Arg162Trp` (not provided), `p.Arg280Trp` (likely benign), `p.Ile207Leu` (benign/-likely benign), and `p.Lys304Glu` (benign). All subsequent steps were identical.

### 5.5 Validation of autosomal dominant estimates using *NFKB1*

To validate our genome-wide probability estimates in an AD gene, we focused on *NFKB1*. Our goal was to compare the expected number of *NFKB1*-related Common Variable Immunodeficiency (CVID) cases, as predicted by our framework, with the reported case count in a well-characterised national-scale PID cohort.

#### 1. Reference dataset

We used a reference dataset reported by Tuijnenburg et al. (39) to build a validation model in an AD disease gene. This study performed whole-genome sequencing of 846 predominantly sporadic, unrelated PID cases from the NIHR BioResource-Rare Diseases cohort. There were 390 CVID cases in the cohort. The study identified *NFKB1* as one of the genes most strongly associated with PID. Sixteen novel heterozygous variants including truncating, missense, and gene deletion variants, were found in *NFKB1* among the CVID cases.

#### 2. Cohort prevalence calculation

Within the cohort, 16 out of 390 CVID cases were attributable to *NFKB1*. Thus, the observed cohort prevalence was 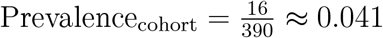 , with a 95% confidence interval (using Wilson’s method) of approximately (0.0254, 0.0656).

#### 3. National estimate based on literature

Based on literature (39; 40; 42), the prevalence of CVID in the general population was estimated as 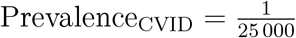. For a UK population of *N*_UK_ ≈ 69 433 632, the expected total number of CVID cases was 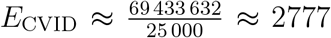. Assuming that the proportion of CVID cases attributable to *NFKB1* is equivalent to the cohort estimate, the literature extrapolated estimate is Estimated NFKB1 cases ≈ 2777 *×* 0.041 ≈ 114, with a median value of approximately 118 and a 95% confidence interval of 70 to 181 cases (derived from posterior sampling).

#### 4. Bayesian adjustment

Recognising that the sequenced cohort cases likely captures the majority of *NFKB1*-related patients (apart from close relatives), but may still miss rare or geographically dispersed variants, we combined the cohort-based and literature-based estimates using two complementary Bayesian approaches:

#### 1. Weighted adjustment (emphasising the cohort, *w* = 0.9)

We assigned 90% weight to the directly observed cohort count (16) and 10% to the extrapolated population estimate (114), thereby accounting, illustratively, for a small fraction of unobserved cases while retaining confidence in our well-characterised cohort: Adjusted Estimate = 0.9 *×* 16 + 0.1 *×* 114 ≈ 26, yielding a 95% CrI of roughly 21 to 33 cases.

#### 2. Mixture adjustment (equal weighting, *w* = 0.5)

To reflect greater uncertainty about how representative the cohort is, we combined cohort and population prevalences equally. We sampled from the posterior distribution of the cohort prevalence, *p* ∼ Beta(16 + 1, 390 − 16 + 1), and mixed this with the literature-based rate at 50% each (39; 40; 42). This yields a median estimate of 67 cases and a wider 95% CrI of approximately 43 to 99 cases, capturing uncertainty in both under-ascertainment and over-generalisation.

#### 5. Predicted total genotype counts

The predicted total synthetic genotype count (before adjustment) was 456, whereas the predicted total genotypes adjusted for `synth_flag` was 0. This higher synthetic count was set based on a minimal risk threshold, ensuring that at least one genotype is assumed to exist (e.g. accounting for a potential unknown de novo variant) even when no variant is observed in gnomAD (as per **section 5.2**).

#### 6. Validation test

Thus, the expected number of *NFKB1*-related CVID cases derived from our genome-wide probability estimates was compared with the observed counts from the UK-based PID cohort. This comparison validates our framework for estimating disease incidence in AD disorders.

### 5.6 Validation study for autosomal recessive CF using *CFTR*

To validate our framework for AR diseases, we focused on Cystic Fibrosis (CF). For comparability sizes between the validation studies, we analysed the most common SNV in the *CFTR* gene, typically reported as `p.Arg117His` (GRCh38 Chr 7:117530975 G/A, MANE Select HGVSp ENST00000003084.11: p.Arg117His). Our goal was to validate our genome-wide probability estimates by comparing the expected number of CF cases attributable to the `p.Arg117His` variant in *CFTR* with the nationally reported case count in a well-characterised disease cohort (43–45).

#### 1. Expected genotype counts

Let *p* denote the allele frequency of the `p.Arg117His` variant and *q* denote the combined frequency of all other pathogenic *CFTR* variants, such that *q* = *P*_tot_ − *p* with *P*_tot_ = ∑_*i*∈CFTR_ *p*_*i*_. Under Hardy-Weinberg equilibrium for an AR trait, the expected frequencies were: *f*_hom_ = *p*^2^ (homozygous for p.Arg117His) and *f*_comphet_ = 2*p q* (compound heterozygotes carrying p.Arg117His and another pathogenic allele). For a population of size *N* (here, *N* ≈ 69 433 632), the expected number of cases were: *E*_hom_ = *N* · *p*^2^, *E*_comphet_ = *N* · 2*p q, E*_total_ = *E*_hom_+ *E*_comphet_.

#### 2. Mortality adjustment

Since CF patients experience increased mortality, we adjusted the expected genotype counts using an exponential survival model (43–45). With an annual mortality rate *λ* ≈ 0.004 and a median age of 22 years, the survival factor was computed as: *S* = exp(−*λ* · 22). Thus, the mortality-adjusted expected genotype count became: *E*_adj_ = *E*_total_ *× S*.

#### 3. Bayesian uncertainty simulation

To incorporate uncertainty in the allele frequency *p*, we modelled *p* as a beta-distributed random variable:

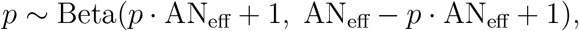

using a large effective allele count (AN_eff_) for illustration. By generating 10,000 posterior samples of *p*, we obtained a distribution of the literature-based adjusted expected counts, *E*_adj_.

#### 4. Bayesian Mixture Adjustment

Since the national registry may not capture all nuances (e.g., reduced penetrance) of *CFTR*-related disease, we further combined the literature-based estimate with the observed national count (714 cases from the UK Cystic Fibrosis Registry 2023 Annual Data Report) using a 50:50 weighting:

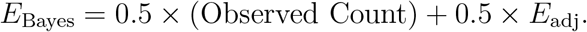

#### 5. Validation test

Thus, the expected number of *CFTR*-related CF cases derived from our genome-wide probability estimates was compared with the observed counts from the UK-based CF registry. This comparison validated our framework for estimating disease incidence in AD disorders.

### 5.7 Validation of SCID-specific estimates using PID-SCID genes

To validate our genome-wide probability estimates for diagnosing a genetic variant in a patient with an PID phenotype, we focused on a subset of genes implicated in SCID. Given that the overall panel corresponds to PID, but SCID represents a rarer subset, the probabilities were converted to values per million PID cases.

#### 1. Incidence conversion

Based on literature, PID occurs in approximately 1 in 1,000 births, whereas SCID occurs in approximately 1 in 100,000 births. Consequently, in a population of 1,000,000 births there are about 1,000 PID cases and 10 SCID cases. To express SCID-related variant counts on a per-million PID scale, the observed SCID counts were multiplied by 100. For example, if a gene is expected to cause SCID in 10 cases within the total PID population, then on a per-million PID basis the count is 10 × 100 = 1,000 cases (across all relevant genes).

#### 2. Prevalence calculation and data adjustment

For each SCID-associated gene (e.g. *IL2RG, RAG1, DCLRE1C*), the observed variant counts in the dataset were adjusted by multiplying by 100 so that the probabilities reflect the expected number of cases per 1,000,000 PID. In this manner, our estimates are directly comparable to known counts from SCID cohorts, rather than to national population counts as in previous validation studies.

#### 3. Integration with prior probability estimates

The predicted genotype occurrence probabilities were derived from our framework across the PID gene panel. These probabilities were then converted to expected case counts per million PID cases by multiplying by 1,000,000. For instance, if the probability of observing a pathogenic variant in *IL2RG* is *p*, the expected SCID-related count becomes *p ×* 10^6^. Similar conversions are applied for all relevant SCID genes.

#### 4. Bayesian Uncertainty and Comparison with Observed Data

To address uncertainty in the SCID-specific estimates, a Bayesian uncertainty simulation was performed for each gene to generate a distribution of predicted case counts on a permillion PID scale. The resulting median estimates and 95% CrIs were then compared against known national SCID counts compiled from independent registries. This comparison permuted a direct evaluation of our framework’s accuracy in predicting the occurrence of SCID-associated variants within a PID cohort.

#### 5. Validation Test

Thus, by converting the overall probability estimates to a per-million PID scale, our framework was directly validated against observed counts for SCID.

### 5.8 Protein network and genetic constraint interpretation

A PPI network was constructed using protein interaction data from STRINGdb (20). We previously prepared and reported on this dataset consisting of 19,566 proteins and 505,968 interactions (https://github.com/DylanLawless/ProteoMCLustR). Node attributes were derived from log-transformed score-positive-total values, which informed both node size and colour. Top-scoring nodes (top 15 based on score) were labelled to highlight prominent interactions. To evaluate group differences in score-positive-total across major disease categories, one-way ANOVA was performed followed by Tukey HSD post hoc tests (and non-parametric Dunn’s test for confirmation). GnomAD v4.1 constraint metrics data was used for the PPI analysis and was sourced from Karczewski et al. (7). This provided transcript-level metrics, such as observed/-expected ratios, LOEUF, Probability of being Loss-of-function Intolerant (pLI), and Z-scores, quantifying LOF and missense intolerance, along with confidence intervals and related annotations for 211,523 observations.

### 5.9 Gene set enrichment test

To test for overrepresentation of biological functions, the prioritised genes were compared against gene sets from MsigDB (including hallmark, positional, curated, motif, computational, GO, oncogenic, and immunologic signatures) and WikiPathways using hypergeometric tests with FUMA (21; 46). The background set consisted of 24,304 genes. Multiple testing correction was applied per data source using the Benjamini-Hochberg method, and gene sets with an adjusted P-value ≤ 0.05 and more than one overlapping gene are reported.

### 5.10 Deriving data-driven classifications of PID genes

To enable reproducible, data-driven grouping of IEI disease genes, we integrated curated clinical annotations with molecular network features. The goal was to formalise immunophenotypic descriptors into a structured format suitable for computational classification, complementing traditional IUIS categories. We recategorised 315 immunophenotypic features from the original IUIS IEI annotations, reducing the original multi-level descriptors (e.g. “decreased CD8, normal or decreased CD4”) first to minimal labels (e.g.”low”) and second to binary outcomes (normal vs. not-normal) for T cells, B cells, neutrophils, and immunoglobulins Each gene was mapped to its PPI cluster derived from STRINGdb and UMAP embeddings from previous steps. We first tested for non-random associations between these four binary immunophenotypes and PPI clusters using *χ*^2^ tests. To generate a data-driven PID classification, we trained a decision tree (rpart) to predict PPI cluster membership from the four immunophenotypic features plus the traditional IUIS Major and Subcategory labels. Hyperparameters (complexity parameter = 0.001, minimum split = 10, minimum bucket = 5, maximum depth = 30) were optimised via five-fold cross validation using the caret framework. Terminal node assignments were then relabelled according to each group’s predominant abnormal feature profile.

### 5.11 Probability of observing AlphaMissense pathogenicity

We obtained the subset pathogenicity predictions from AlphaMissense via the AlphaFold database and whole genome data from the studies data repository(12; 37). The AlphaMissense data (genome-aligned and amino acid substitutions) were merged with the panel variants based on genomic coordinate and HGVSc annotation. Occurrence probabilities were log-transformed and adjusted (y-axis displaying log10(occurrence prob + 1e-5) + 5)), to visualise the distribution of pathogenicity scores across the residue sequence. A Kruskal-Wallis test was used to compare the observed disease probability across clinical classification groups.

### 5.12 Probability model definitions

Estimating disease risk requires accounting for both variant penetrance, *P* (*D* | *G*), where *D* denotes the disease state and *G* the genotype, and the fraction of cases attributable to a given variant, *P* (*G* | *D*). In a fully penetrant single-variant model (*P* (*D* | *G*) = 1), the lifetime risk *P* (*D*) equals the genotype frequency *P* (*G*). For an allele with population frequency *p*, this gives *P* (*D*) = *p*^2^ for a recessive mode of inheritance and *P* (*D*) = 2*p*(1 − *p*) ≈ 2*p* for a dominant mode. With incomplete penetrance, *P* (*D*) = *P* (*G*) *P* (*D* | *G*), and when multiple variants contribute to disease,

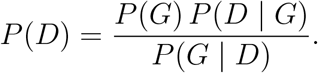

Because both *P* (*D* | *G*) and *P* (*G* | *D*) are often uncertain, we integrate ClinVar clinical classifications, population allele frequencies and curated gene-disease associations, assuming James-Stein shrinkage to derive robust aggregate priors. By focusing on a filtered set of variants 𝒱 where each *P* (*G*_*i*_ | *D*) is the probability that disease *D* is attributable to variant *i* and assuming _*i*∈𝒱_ *P* (*G*_*i*_ | *D*) ≈ 1, we obtain calibrated estimates of genotype frequency *P* (*G*) despite uncertainty in individual parameters.

## Acronyms

ACMG: American College of Medical Genetics and Genomics
ACAT: Aggregated Cauchy Association Test
AD: Autosomal Dominant
AF: Allele Frequency
ANOVA: Analysis of Variance
AR: Autosomal Recessive
BMF: Bone Marrow Failure
CD: Complement Deficiencies
CI: Confidence Interval
CrI: Credible Interval
CF: Cystic Fibrosis
*CFTR*: Cystic Fibrosis Transmembrane Conductance Regulator
CVID: Common Variable Immunodeficiency
*DCLRE1C*: DNA Cross-Link Repair 1C
dbNSFP: database for Non-Synonymous Functional Predictions
GE: Genomics England
gnomAD: Genome Aggregation Database
gVCF: genomic variant call format
HGVS: Human Genome Variation Society
HPC: High-Performance Computing
HSD: Honestly Significant Difference
HWE: Hardy-Weinberg Equilibrium
IEI: Inborn Errors of Immunity
Ig: Immunoglobulin
*IL2RG*: Interleukin 2 Receptor Subunit Gamma
InDel: Insertion/Deletion
IUIS: International Union of Immunological Societies
LD: Linkage Disequilibrium
LOEUF: Loss-Of-function Observed/Expected Upper bound Fraction
LOF: Loss-of-Function
MOI: Mode of Inheritance
*NFKB1*: Nuclear Factor Kappa B Subunit 1
OMIM: Online Mendelian Inheritance in Man
PID: Primary Immunodeficiency
PPI: Protein-Protein Interaction
pLI: Probability of being Loss-of-function Intolerant
QC: Quality Control
*RAG1*: Recombination activating gene 1
SCID: Severe Combined Immunodeficiency
SNV: Single Nucleotide Variant
SKAT: Sequence Kernel Association Test
STRINGdb: Search Tool for the Retrieval of Interacting Genes/Proteins
TP: true positive
FP: false positive
TN: true negative
FN: false negative
*TNFAIP3*: Tumor necrosis factor, alpha-induced protein 3
UMAP: Uniform Manifold Approximation and Projection
UniProt: Universal Protein Resource
VCF: variant call format
VEP: Variant Effect Predictor
VRE: variant risk estimate
XL: X-Linked

## 6 Supplemental

Supplemental data are presented under the same headings that correspond to their relevant main text sections.

### 6.1 Occurrence probability across disease genes

### 6.2 Integrating observed true positives and unobserved false negatives into a single, actionable conclusion

**Table S1:**
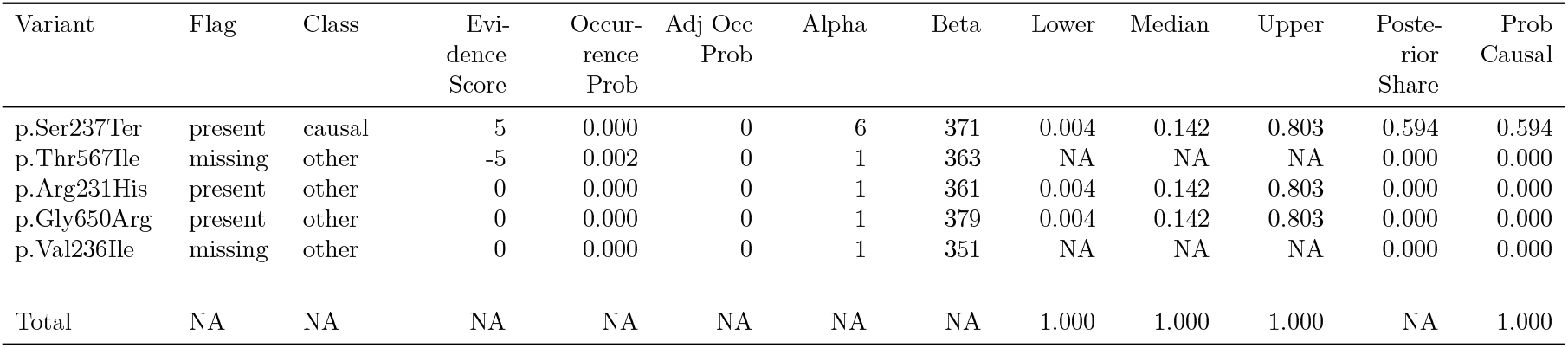
Result of clinical genetics diagnosis scenario 1 including metadata. The most strongly supported observed variant was `p.Ser237Ter` (posterior: 0.594). The strongest unsequenced variant was `p.Thr567Ile` (posterior: 0). The total probability of a causal diagnosis given the available evidence was 1 (95% CI: 1–1).

| Variant | Flag | Class | Evidence Score | Occurrence Prob | Adj Occ Prob | Alpha | Beta | Lower | Median | Upper | Posterior Share | Prob Causal |
| --- | --- | --- | --- | --- | --- | --- | --- | --- | --- | --- | --- | --- |
| p.Ser237Ter | present | causal | 5 | 0.000 | 0 | 6 | 371 | 0.004 | 0.142 | 0.803 | 0.594 | 0.594 |
| p.Thr567Ile | missing | other | -5 | 0.002 | 0 | 1 | 363 | NA | NA | NA | 0.000 | 0.000 |
| p.Arg231His | present | other | 0 | 0.000 | 0 | 1 | 361 | 0.004 | 0.142 | 0.803 | 0.000 | 0.000 |
| p.Gly650Arg | present | other | 0 | 0.000 | 0 | 1 | 379 | 0.004 | 0.142 | 0.803 | 0.000 | 0.000 |
| p.Val236Ile | missing | other | 0 | 0.000 | 0 | 1 | 351 | NA | NA | NA | 0.000 | 0.000 |
| Total | NA | NA | NA | NA | NA | NA | NA | 1.000 | 1.000 | 1.000 | NA | 1.000 |

**Table S2:** Result of clinical genetics diagnosis scenario 2 including metadata. The most strongly supported observed variant was `p.Ser237Ter` (posterior: 0.381). The strongest unsequenced variant was `c.159+1G>A` (posterior: 0.353). The total probability of a causal diagnosis given the available evidence was 0.52 (95% CI: 0.248–0.787).

| Variant | Flag | Class | Evidence Score | Occurrence Prob | Adj Occ Prob | Alpha | Beta | Lower | Median | Upper | Posterior Share | Prob Causal |
| --- | --- | --- | --- | --- | --- | --- | --- | --- | --- | --- | --- | --- |
| p.Ser237Ter | present | causal | 5.0 | 0.000 | 0 | 6.0 | 371 | 0.003 | 0.096 | 0.557 | 0.381 | 0.381 |
| c.159+1G>A | missing | causal | 4.5 | 0.000 | 0 | 5.5 | 367 | NA | NA | NA | 0.353 | 0.353 |
| p.Thr567Ile | missing | other | -5.0 | 0.002 | 0 | 1.0 | 365 | NA | NA | NA | 0.000 | 0.000 |
| p.Arg231His | present | other | 0.0 | 0.000 | 0 | 1.0 | 359 | 0.003 | 0.096 | 0.557 | 0.000 | 0.000 |
| p.Gly650Arg | present | other | 0.0 | 0.000 | 0 | 1.0 | 349 | 0.003 | 0.096 | 0.557 | 0.000 | 0.000 |
| p.Val236Ile | missing | other | 0.0 | 0.000 | 0 | 1.0 | 363 | NA | NA | NA | 0.000 | 0.000 |
| Total | NA | NA | NA | NA | NA | NA | NA | 0.248 | 0.520 | 0.787 | NA | 0.520 |

**Table S3:** Result of clinical genetics diagnosis scenario 3 including metadata. No observed variants were detected in this scenario. The strongest unsequenced variant was `p.Cys243Arg` (posterior: 0.366). The total probability of a causal diagnosis given the available evidence was 0 (95% CI: 0–0).

| Variant | Flag | Class | Evidence Score | Occurrence Prob | Adj Occ Prob | Alpha | Beta | Lower | Median | Upper | Posterior Share | Prob Causal |
| --- | --- | --- | --- | --- | --- | --- | --- | --- | --- | --- | --- | --- |
| p.Cys243Arg | missing | causal | 5.0 | 0.000 | 0.000 | 6 | 341 | NA | NA | NA | 0.366 | 0.366 |
| p.Tyr246Ter | missing | causal | 4.0 | 0.000 | 0.000 | 5 | 369 | NA | NA | NA | 0.284 | 0.284 |
| p.Lys304Glu | missing | other | -5.0 | 0.000 | 0.000 | 1 | 353 | NA | NA | NA | 0.000 | 0.000 |
| p.Ile207Leu | missing | other | -4.5 | 0.000 | 0.000 | 1 | 359 | NA | NA | NA | 0.000 | 0.000 |
| p.His646Pro | missing | other | 0.0 | 0.002 | 0.001 | 1 | 377 | NA | NA | NA | 0.000 | 0.000 |
| p.Arg280Trp | missing | other | -4.0 | 0.000 | 0.000 | 1 | 357 | NA | NA | NA | 0.000 | 0.000 |
| p.Thr635Ile | missing | other | 0.0 | 0.000 | 0.000 | 1 | 349 | NA | NA | NA | 0.000 | 0.000 |
| p.Arg162Trp | missing | other | 0.0 | 0.000 | 0.000 | 1 | 369 | NA | NA | NA | 0.000 | 0.000 |
| Total | NA | NA | NA | NA | NA | NA | NA | 0 | 0 | 0 | NA | 0.000 |

**Figure S1:**
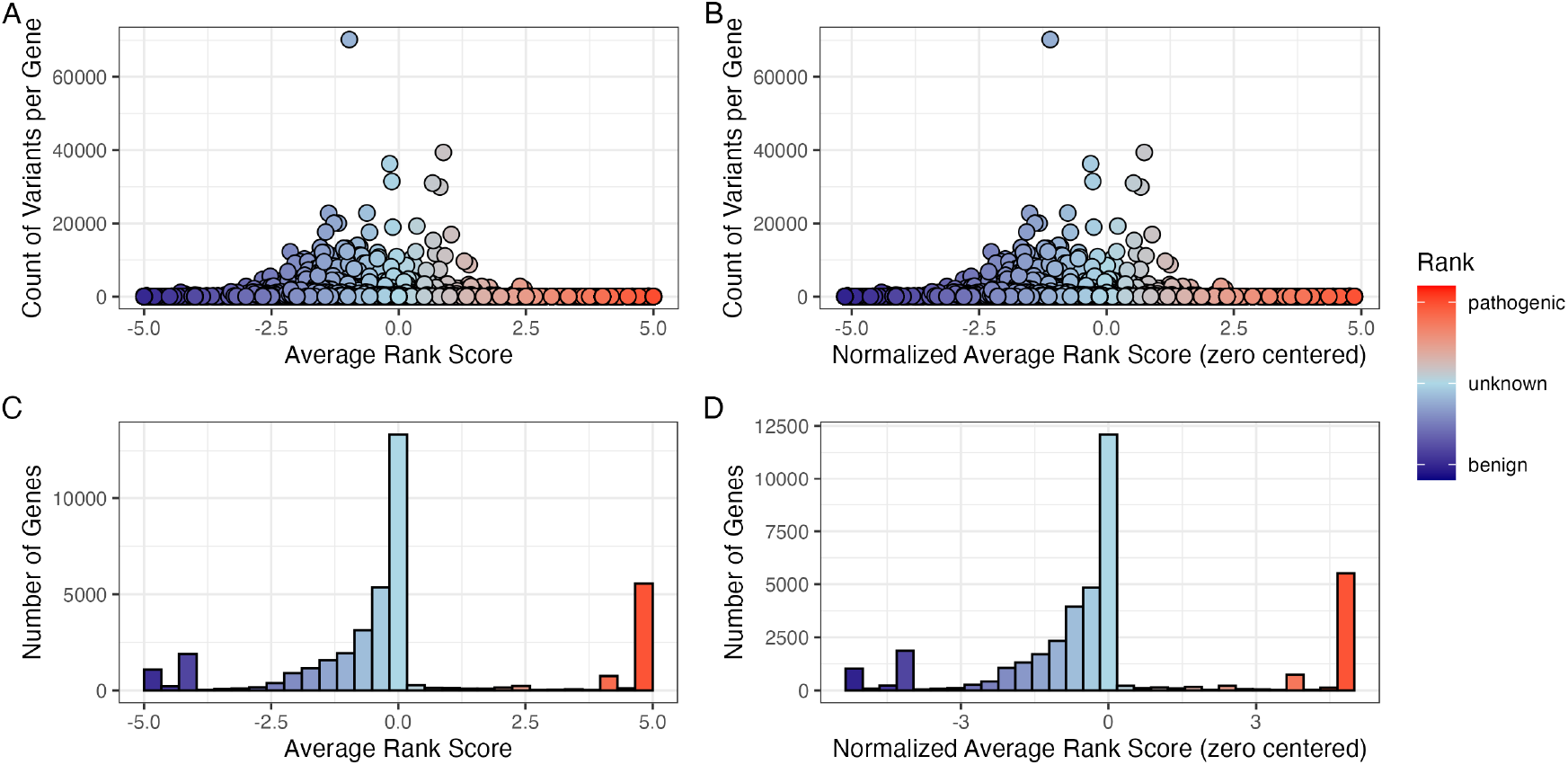
Global distribution of ClinVar clinical-significance classification scoring. (A) Number of variants per gene containing the assigned score for each ClinVar classification term (−5 to +5). (B) The same data after normalisation by zero centring the average rank score. (C) The tally of genes for their average rank and (D) after normalisation. No normalisation was required for the scoring system as shown by comparison of A-C and B-D.

**Figure S2:**
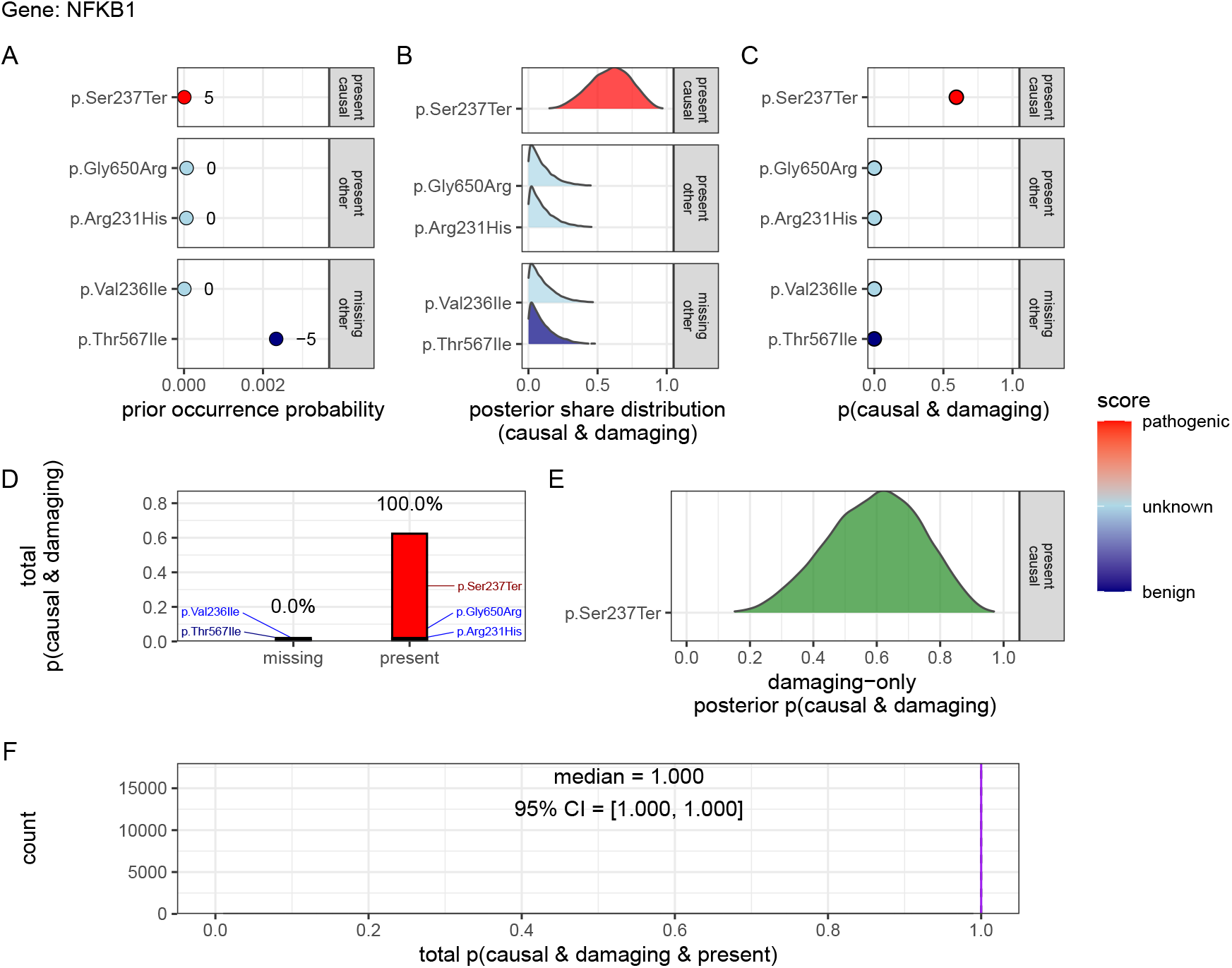
Quantification of present (TP) and no missing (FN) causal genetic variants for disease in *NFKB1* (scenario 1) Only one known pathogenic variant, `p.Ser237Ter` , was observed and all previously reported pathogenic positions were successfully sequenced and confirmed as reference (true negatives). Panels (A-F) follow the same structure as scenario 2 described in **Figure 2**, culminating in a gene-level posterior probability of 1 (95 % CrI: 0.99-1.00), with full support assigned to the observed allele given the available evidence. Pathogenicity scores (-5 to +5) are annotated.

**Figure S3:**
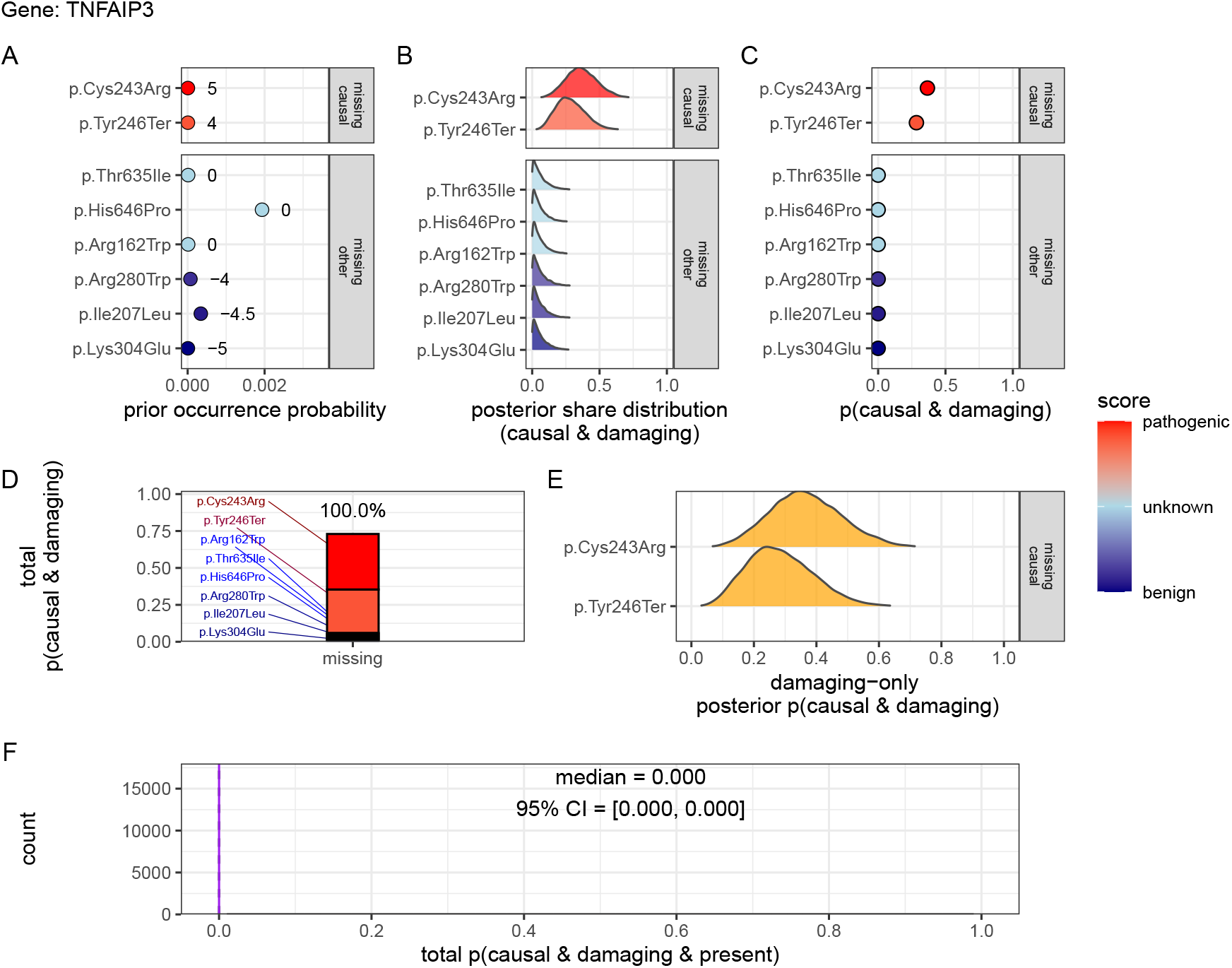
Quantification of no present (TP) in *NFKB1* and only missing (FN) causal genetic variants for disease in *TNFAIP3* (scenario 3). No known causal variants were observed in *NFKB1*, but one representative unsequenced allele was selected from each distinct ClinVar classification and treated as a potential false negative. Panels (A-F) follow the same structure as scenario 2 described in **Figure 2**. The posterior reflects uncertainty across multiple plausible but unobserved variants, resulting in low CrI (0-0) and 100% missing overall attribution in contrast to scenarios where known pathogenic variants were observed. For this patient, we have no evidence of a causal variant since the only top candidates are not yet accounted for. Pathogenicity scores (-5 to +5) are annotated in (A).

### 6.3 Validation studies

#### 6.3.1 Validation of dominant disease occurrence with *NFKB1*

To validate our genome-wide probability estimates for AD disorders, we focused on *NFKB1*. We used a reference dataset from Tuijnenburg et al. (39), in which whole-genome sequencing of 846 PID patients identified *NFKB1* as one of the genes most strongly associated with the disease, with 16 *NFKB1*-related CVID cases attributed to AD heterozygous variants. Our goal was to compare the predicted number of *NFKB1*-related CVID cases with the reported count in this well-characterised national-scale cohort.

Our model calculated 0 known pathogenic variant *NFKB1*-related CVID cases in the UK with a minimal risk of 456 unknown de novo variants. In the reference cohort, 16 *NFKB1* CVID cases were reported. We additionally wanted to account for potential under-reporting in the reference study. We used an extrapolated national CVID prevalence which yielded a median estimate of 118 cases (95% CI: 70-181), while a Bayesian-adjusted mixture estimate produced a median of 67 cases (95% CI: 43-99). **Figure S4 (A)** illustrates that our predicted values reflect these ranges and are closer to the observed count. This case supports the validity of our integrated probability estimation framework for AD disorders, and represents a challenging example where pathogenic SNV are not reported in the reference population of gnomAD. Our min-max values successfully contained the true reported values.

#### 6.3.2 Validation of recessive disease occurrence with *CFTR*

Our analysis predicted the number of CF cases attributable to carriage of the `p.Arg117His` variant (either as homozygous or as compound heterozygous with another pathogenic allele) in the UK. Based on HWE calculations and mortality adjustments, we predicted approximately 648 cases arising from biallelic variants and 160 cases from homozygous variants, resulting in a total of 808 expected cases.

In contrast, the nationally reported number of CF cases was 714, as recorded in the UK Cystic Fibrosis Registry 2023 Annual Data Report (43). To account for factors such as reduced penetrance and the mortality-adjusted expected genotype, we derived a Bayesian-adjusted estimate via posterior simulation. Our Bayesian approach yielded a median estimate of 740 cases (95% CI: 696, 786) and a mixture-based estimate of 727 cases (95% CI: 705, 750). **Figure S4 (B)** illustrates the close concordance between the predicted values, the Bayesian-adjusted estimates, and the national report supports the validity of our approach for estimating disease.

**Figure S5** shows the final values for these genes of interest in a given population size and phenotype. It reveals that an allele frequency threshold of approximately 0.000007 is required to observe a single heterozygous carrier of a disease-causing variant in the UK population for both genes. However, owing to the AR MOI pattern of *CFTR*, this threshold translates into more than 100,000 heterozygous carriers, compared to only 456 carriers for the AD gene *NFKB1*. Note that this allele frequency threshold, being derived from the current reference population, represents a lower bound that can become more precise as public datasets continue to grow. This marked difference underscores the significant impact of MOI patterns on population carrier frequencies and the observed disease prevalence.

#### 6.3.3 Interpretation of ClinVar variant occurrences

**Figure S6** shows the two validation study PID genes, representing AR and dominant MOI. **Figure S6 (A)** illustrates the overall probability of an affected birth by ClinVar variant classification, whereas **Figure S6 (B)** depicts the total expected number of cases per classification for an example population, here the UK, of approximately 69.4 million.

#### 6.3.4 Validation of SCID-specific disease occurrence

Given that SCID is a subset of PID, our probability estimates reflect the likelihood of observing a genetic variant as a diagnosis when the phenotype is PID. However, we additionally tested our results against SCID cohorts in **Figure S8**. The summarised raw cohort data for SCID-specific gene counts are summarised and compared across countries in **Figure S7**. True counts for *IL2RG* and *DCLRE1C* from ten distinct locations yielded 95% CI surrounding our predicted values. For *IL2RG*, the prediction was low (approximately 1 case per 1,000,000 PID), as expected since loss-of-function variants in this XL gene are highly deleterious and rarely observed in gnomAD. In contrast, the predicted value for *RAG1* was substantially higher (553 cases per 1,000,000 PID) than the observed counts (ranging from 0 to 200). We attributed this discrepancy to the lower penetrance and higher background frequency of *RAG1* variants in recessive inheritance, whereby reference studies may underreport the true national incidence. Overall, we report that agreement within an order of magnitude was tolerable given the inherent uncertainties from reference studies arising from variable penetrance and allele frequencies.

**Figure S4:**
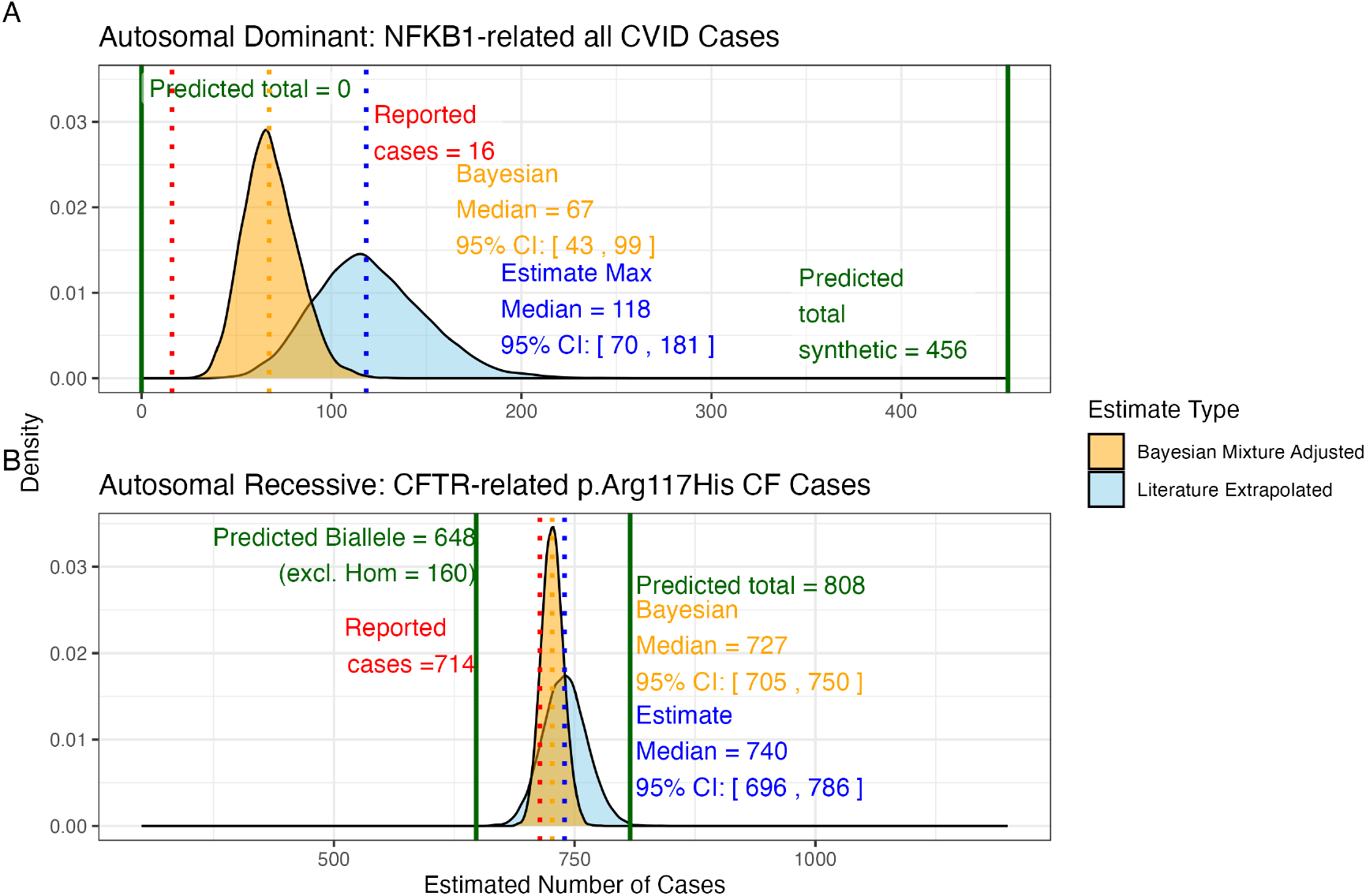
Prior probabilities compared to validation disease cohort metrics. (A) Density distributions for the number of *NFKB1*-related CVID cases in the UK. Our model (green) predicted 456 cases, which falls between the observed cohort count (red) of 390 and the upper extrapolated values. The blue curve represents maximum count of 1280, and the orange curve shows the Bayesian-adjusted mixture estimate of 835. (B) Density distributions for *CFTR*-related `p.Arg117His` CF cases. Our model (green) predicted 648 biallelic cases and 808 total cases. The nationally reported case count (red) was 714. The blue curve represents maximum extrapolated count of 740, and the orange curve shows the Bayesian-adjusted mixture estimate of 727. We observed close agreement among the reported disease cases and our integrated probability estimation framework.

**Figure S5:**
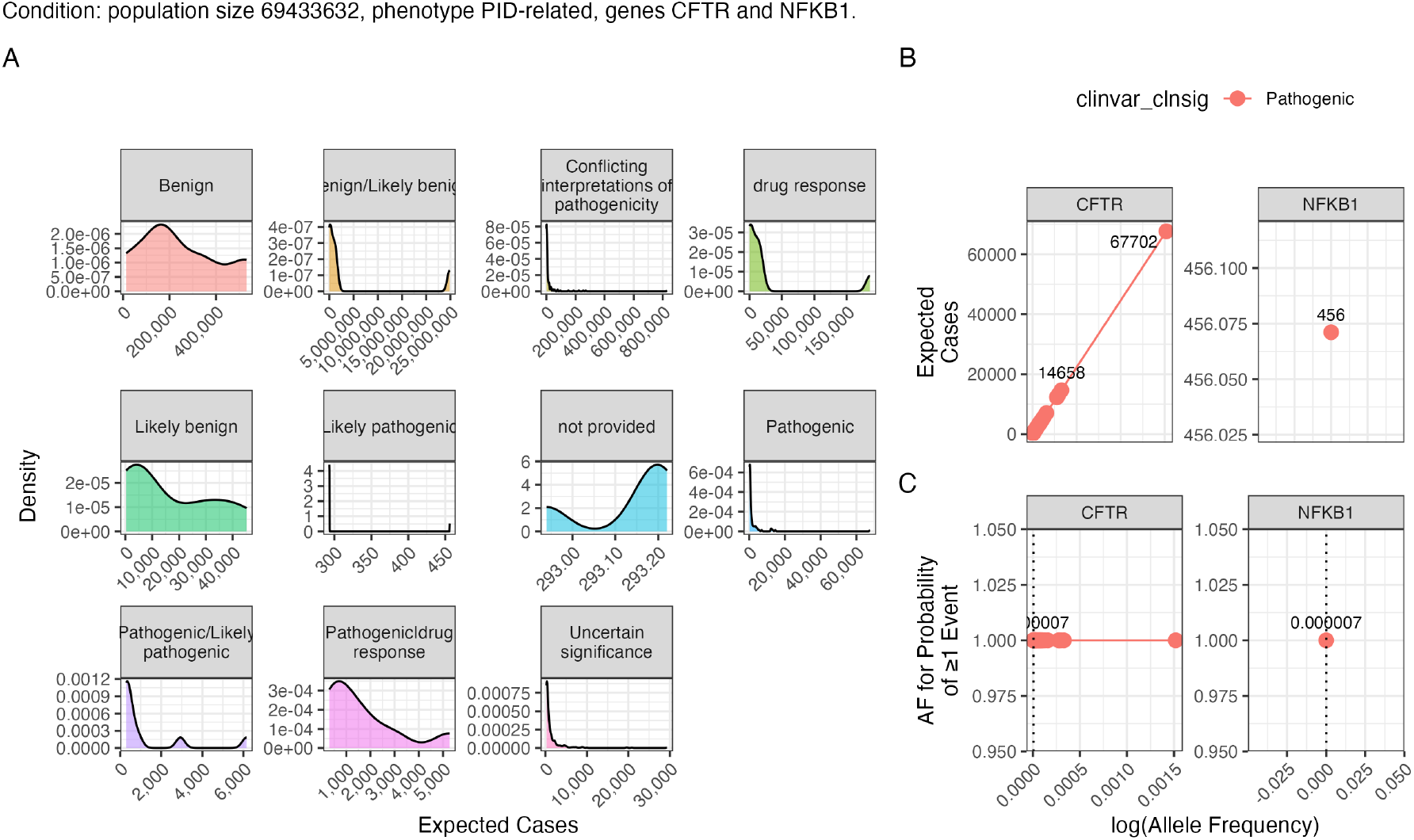
Interpretation of probability of observing a variant classification. The result from the chosen validation genes *CFTR* and *NFKB1* are shown. Case counts are dependant on the population size and phenotype. (A) The density plots of expected observations by ClinVar clinical significance. We then highlight the values for pathogenic variants specifically showing; (B) the allele frequency versus expected cases in this population size and (C) the probability of observing at least one event in this population size.

**Figure S6:**
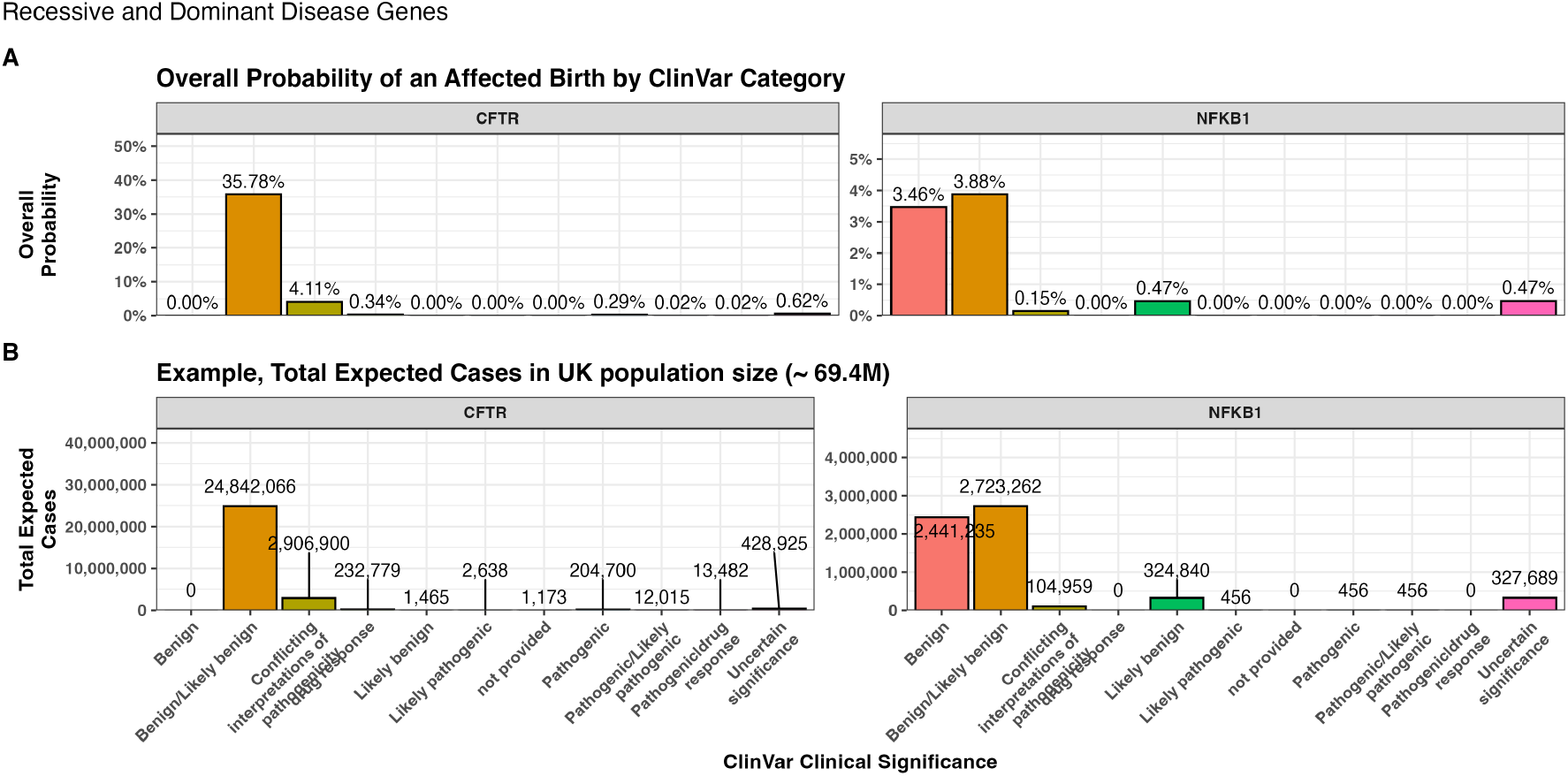
Combined bar charts summarising the genome-wide analysis of ClinVar clinical significance for the PID gene panel. Panel (A) shows the overall probability of an affected birth by variant classification, and (B) displays the total expected number of cases per classification, both stratified by gene. These integrated results illustrate the variability in variant observations across genes and underpin our validation of the probability estimation framework.

**Figure S7:**
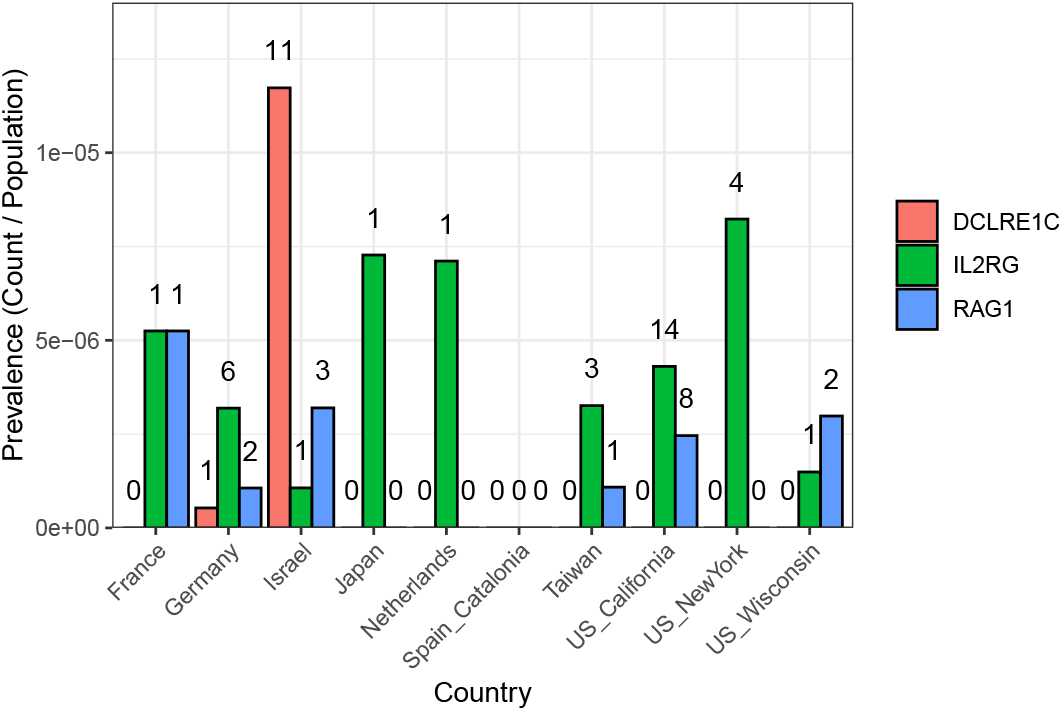
SCID-specific gene comparison across regions. The bar plot shows the prevalence of SCID-related cases (count divided by population) for each gene and country (or region), with numbers printed above the bars representing the actual counts in the original cohort (ranging from 0 to 11 per region and gene).

**Figure S8:**
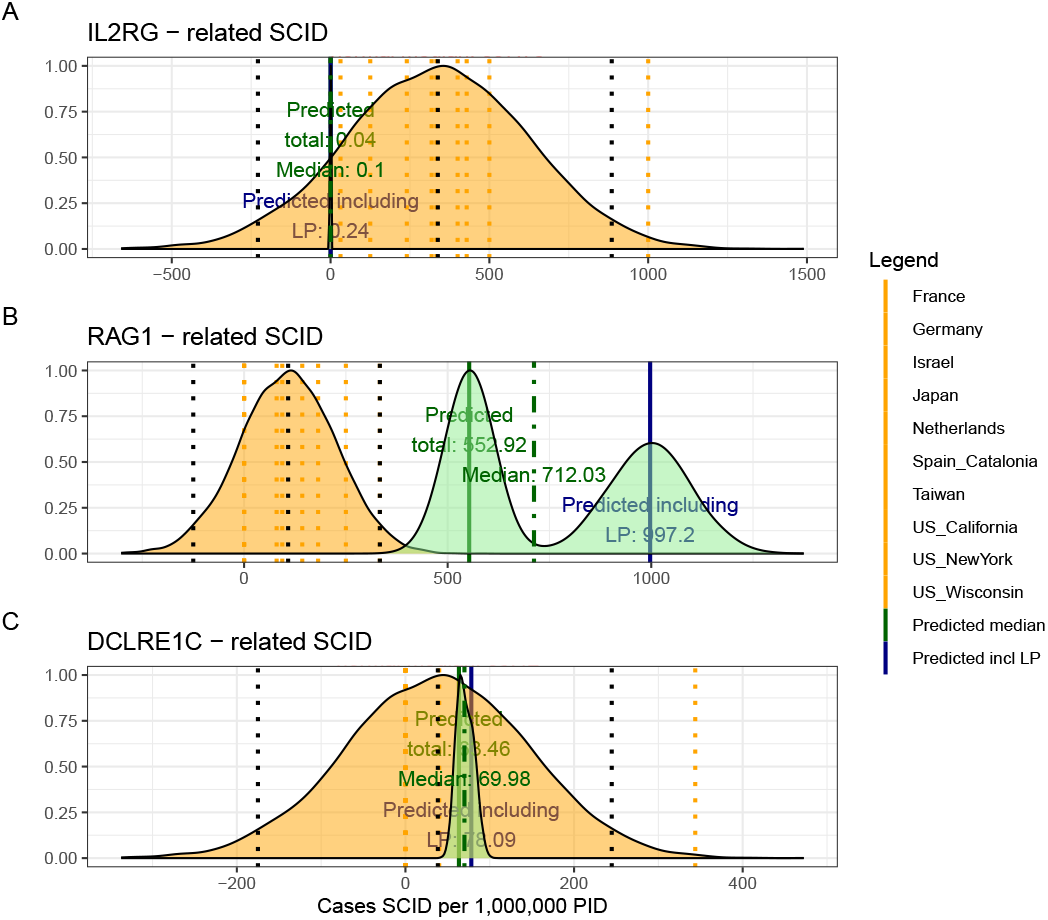
Combined SCID-specific predictions and observed rates per 1,000,000 PID. The figure presents density distributions for the predicted SCID case counts (per 1,000,000 PID) for three genes: *IL2RG, RAG1*, and *DCLRE1C*. Country-specific rates (displayed as dotted vertical lines) are overlaid with the overall predicted distributions for pathogenic and likely pathogenic variants (solid lines with annotated medians). For *IL2RG*, the low predicted value is consistent with the high deleteriousness of loss-of-function variants in this X-linked gene, while *RAG1* exhibits considerably higher predicted counts, reflecting its lower penetrance in an autosomal recessive context.

### 6.4 Quantifying variant probabilities improves classification of inborn errors of immunity

#### 6.4.1 Genes with high pathogenic variant burden show structured network constraint

**Figure S9:**
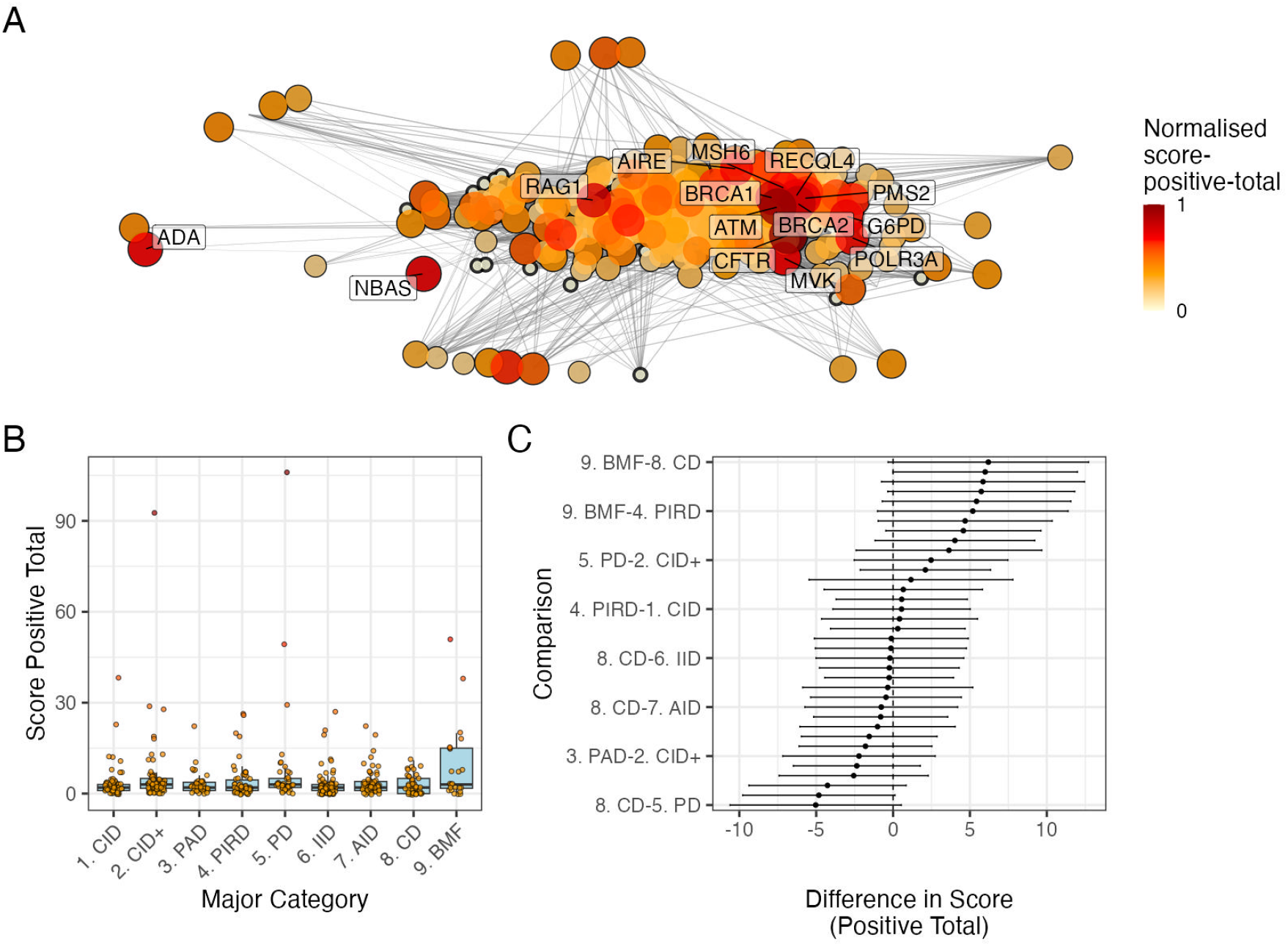
PPI network and score-positive-total ClinVar significance variants. (A) PPI network of disease-associated genes. Node size and colour represent the log-transformed score-positive-total, the top 15 genes/proteins with the highest probability of being observed in disease are labelled. (B) Distribution of score-positive-total across the major IEI disease categories. (C) Tukey HSD comparisons of mean differences in score-positive-total among all pairwise disease categories. Every 5th label is shown on y-axis.

#### 6.4.2 Constrained subnetworks reveal distinct functional disease signatures

**Figure S10:**
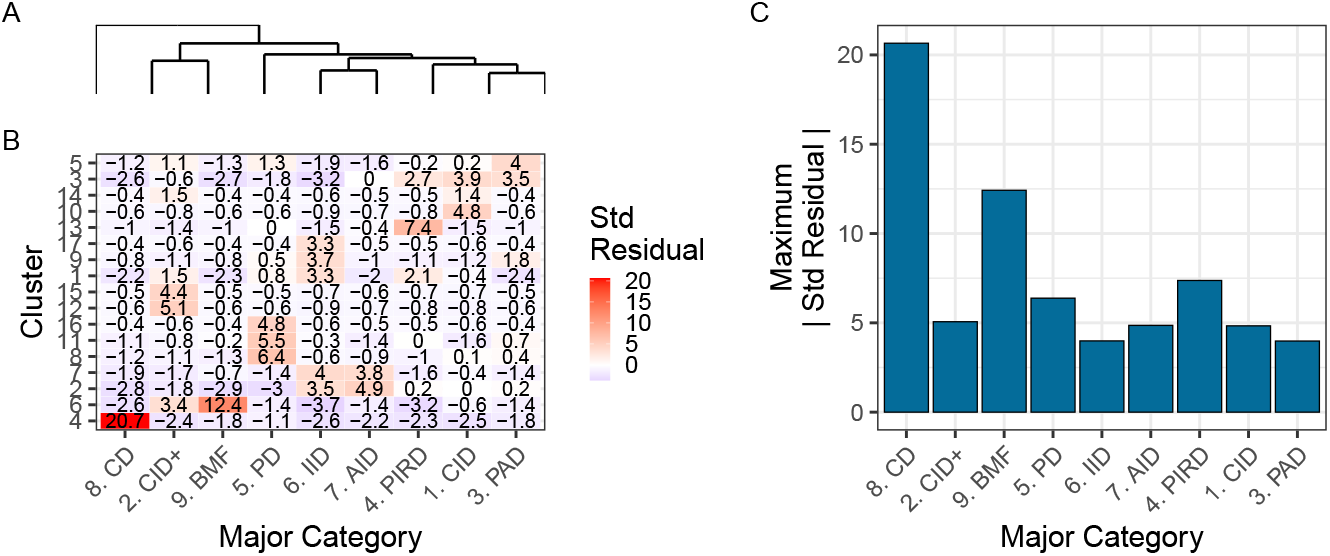
Hierarchical clustering of enrichment scores. The heatmap displays standardised residuals for major disease categories (x-axis) across network clusters (y-axis). A dendrogram groups similar disease categories, and the bar plot shows the maximum absolute residual per category. (8) CD and (9)BMF show the highest vales, indicating significant enrichment or depletion (residuals > |2|). Definitions in **Box 1**.

**Figure S11:**
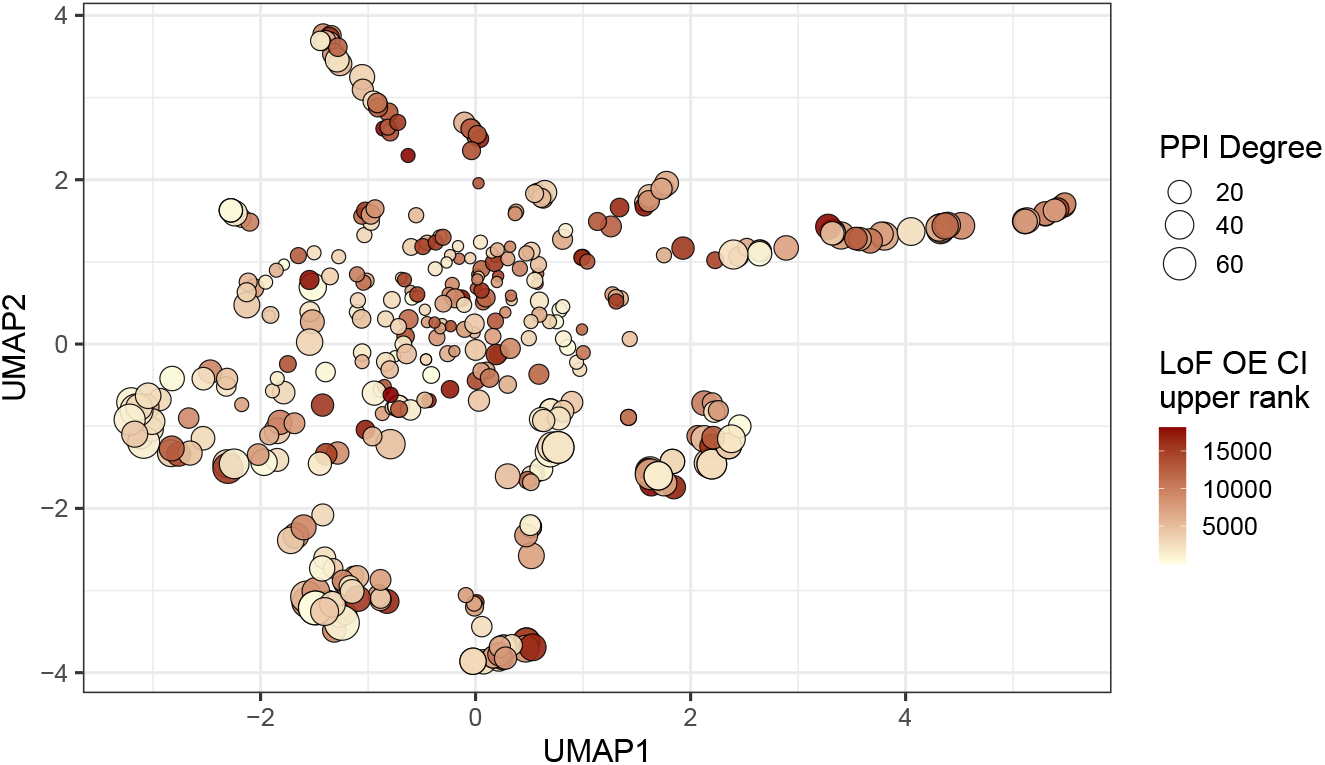
Analysis of PPI degree versus LOEUF upper rank with UMAP embedding of the PPI network. The relationship between PPI degree (size) and LOEUF upper rank (color) across gene clusters. No clear patterns are evident.

**Figure S12:**
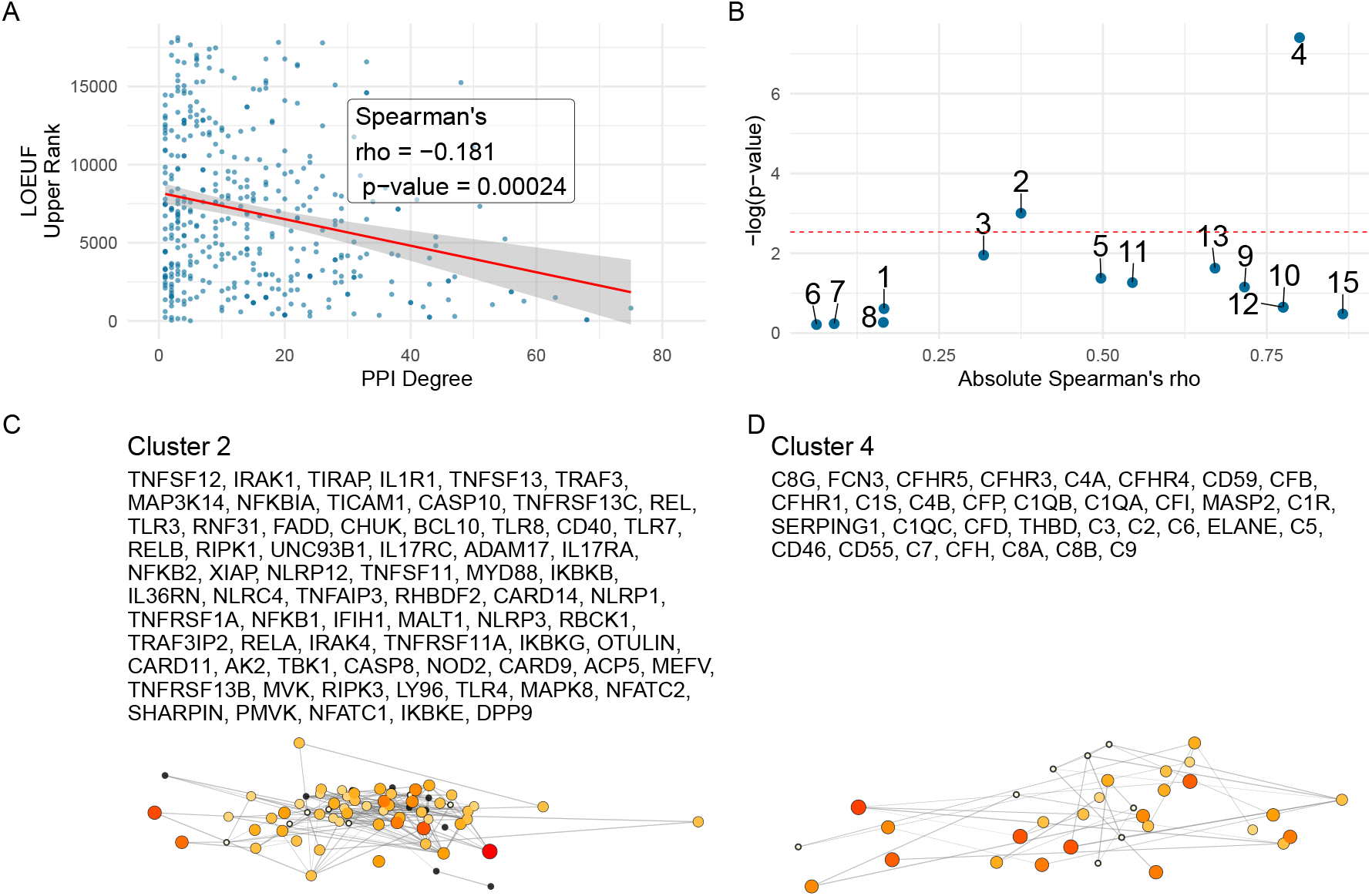
Correlation between PPI degree and LOEUF upper rank. **(A)** Ananlysis across all genes revealed a weak, significant negative correlation between PPI degree and LOEUF upper rank. **(B)** The cluster-wise analysis showed that clusters 2 and 4 exhibited moderate to strong correlations, while other clusters display weak or non-significant relationships. **(C) and (D)** Shows the new network plots for the significantly enriched clusters based on gnomAD constraint metrics.

**Figure S13:**
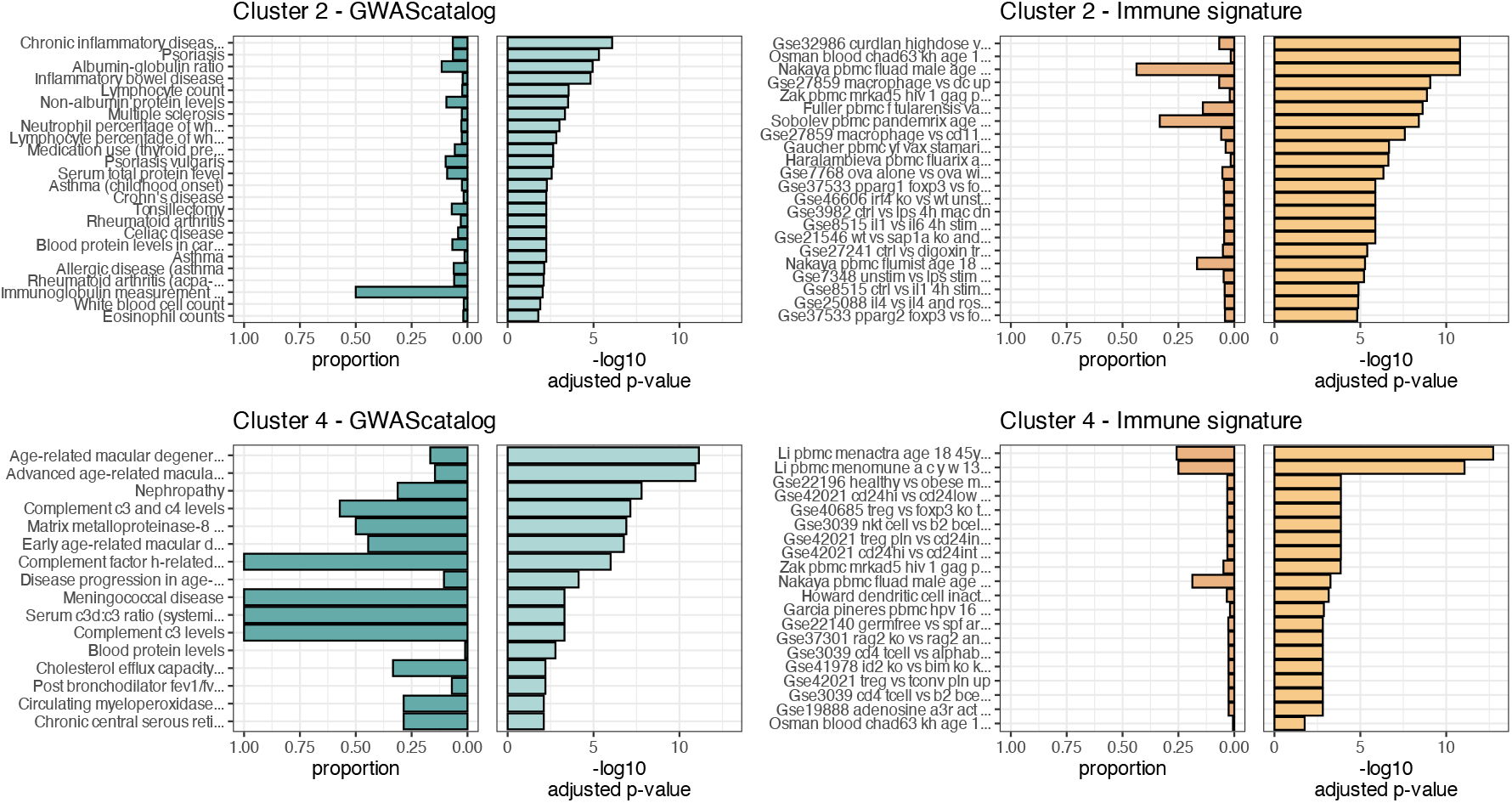
Composite Enrichment Profiles for IEI Gene Sets. We selected the top two enriched clusters (as per **Figure S12**) and performed functional enrichment analysis derived from known disease associations. For each gene set, the left panel displays the proportion of input genes overlapping with a curated gene set, and the right panel shows the − log_10_ adjusted p-value from hypergeometric testing. These profiles, stratified by cluster (Cluster 2 and Cluster 4) and by gene set category (GWAScatalog and Immunologic Signatures), highlight distinct enrichment patterns that reflect differential pathogenic variant loads in the IEI gene panels.

**Figure S14:**
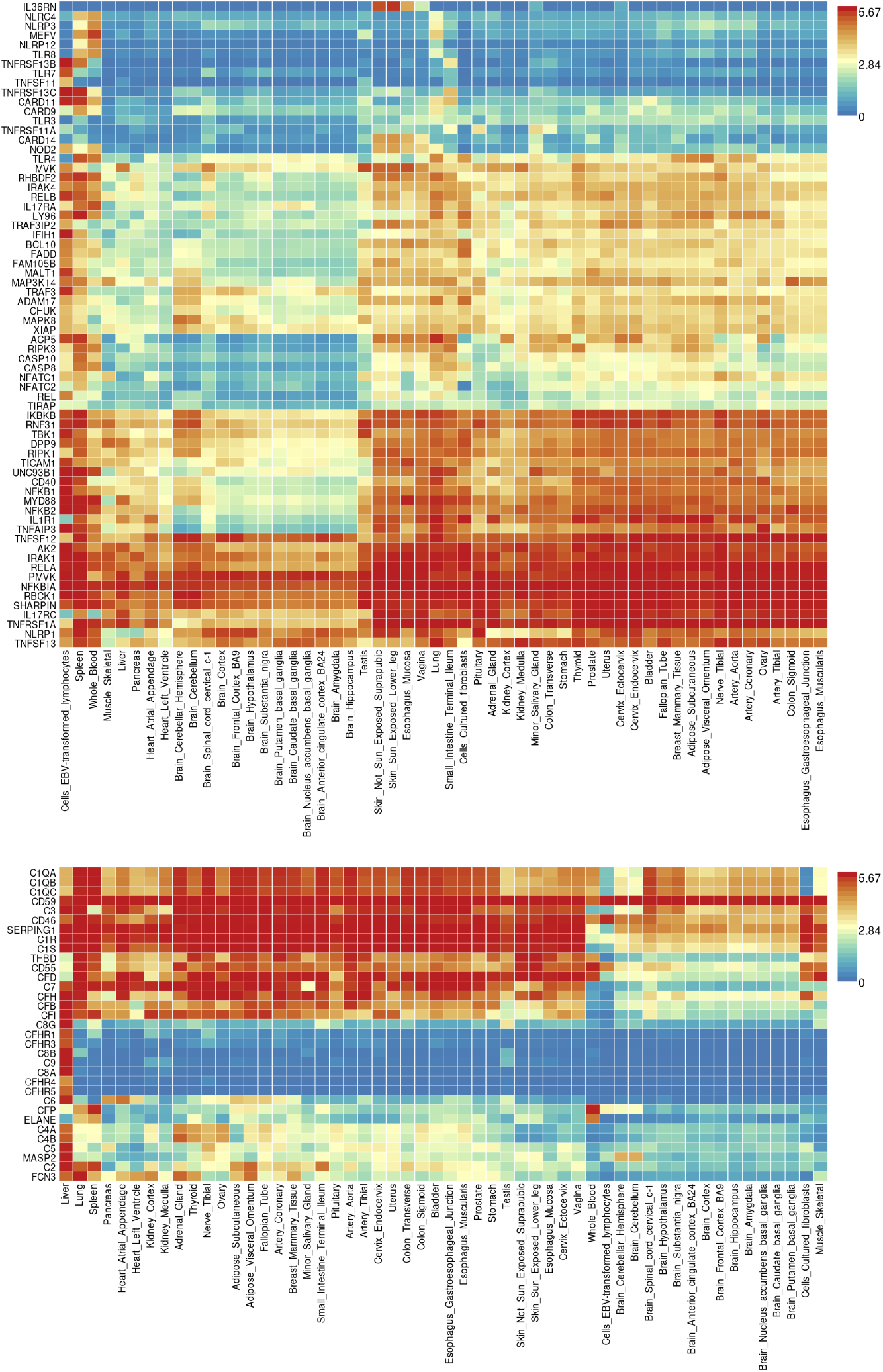
Gene Expression Heatmaps for IEI Genes. GTEx v8 data from 54 tissue types display the average expression per tissue label (log_2_ transformed) for the IEI gene panels. Top: Cluster 2; Bottom: Cluster 4.

#### 6.4.3 Data-driven classifications of PID genes

**Figure S15:**
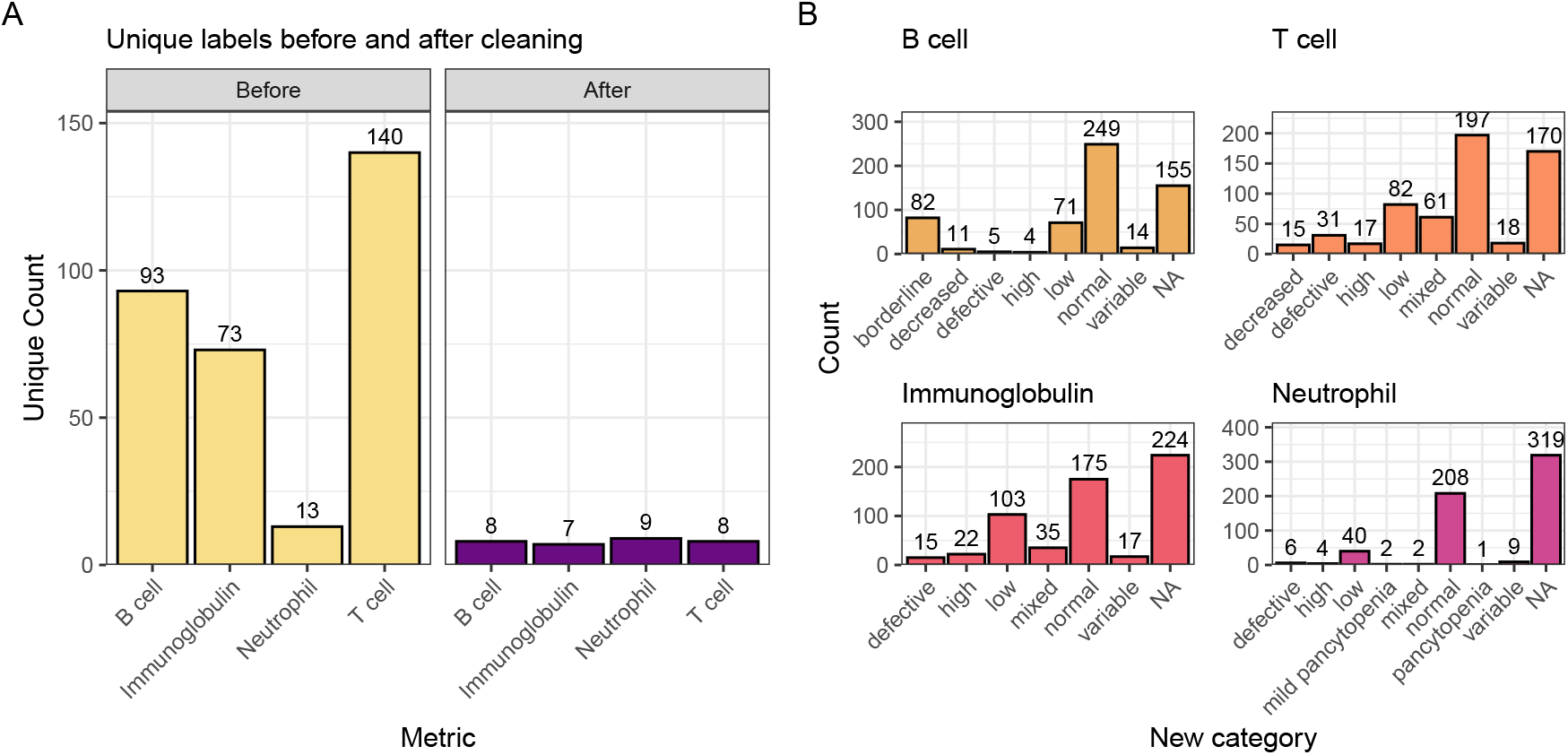
Distribution of immunophenotypic features before and after recategorisation. The original IUIS IEI descriptions contain information such as T cell-related “decreased CD8, normal or decreased CD4 cells” which we recategorise as “low”. The bar plot shows the count of unique labels for each status (normal, not_normal) across the T cell, B cell, Ig, and Neutrophil features.

**Figure S16:**
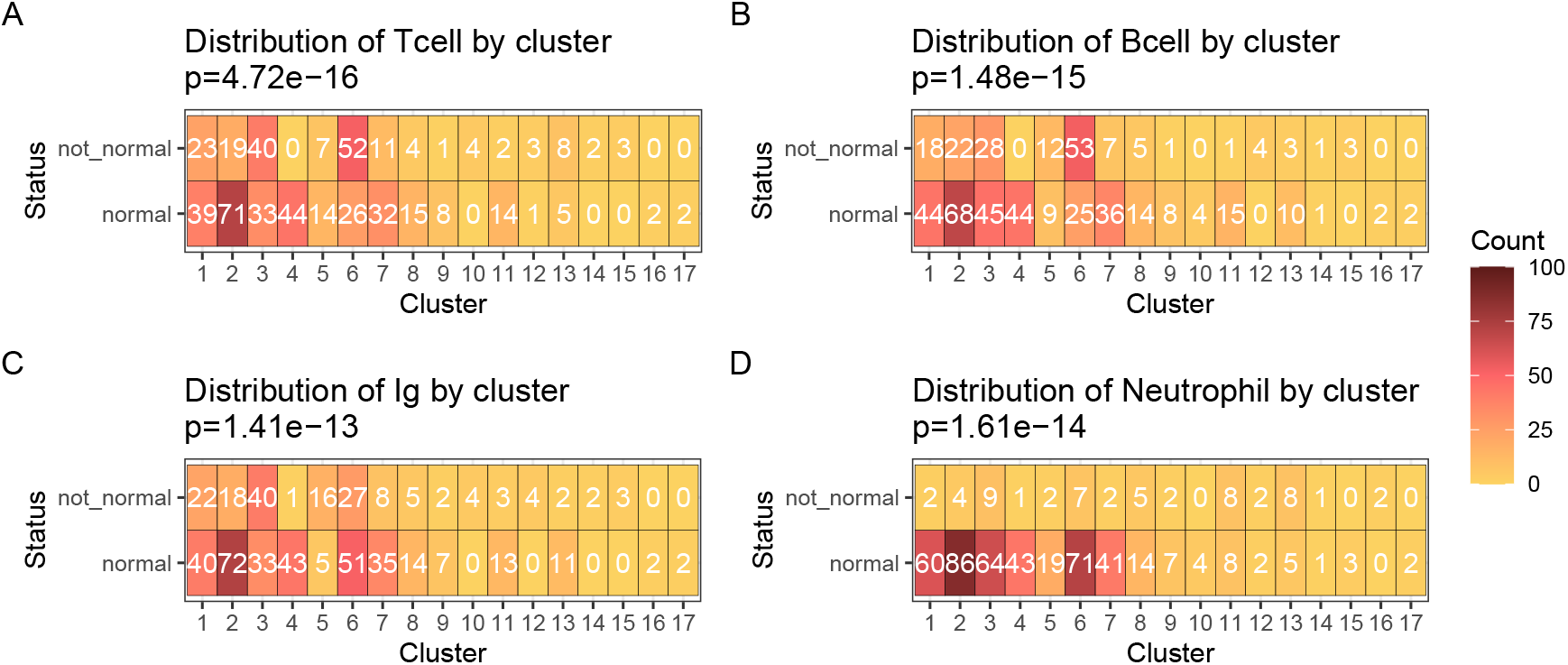
Heatmaps of clinical feature distributions by PPI cluster. The heatmaps display the count of observations for abnormality of each clinical feature (A) T cell, (B) B cell, (C) Immunoglobulin, (D) Neutrophil, in relation to the PPI clusters, with p-values from chi-square tests annotated in the titles.

**Figure S17:**
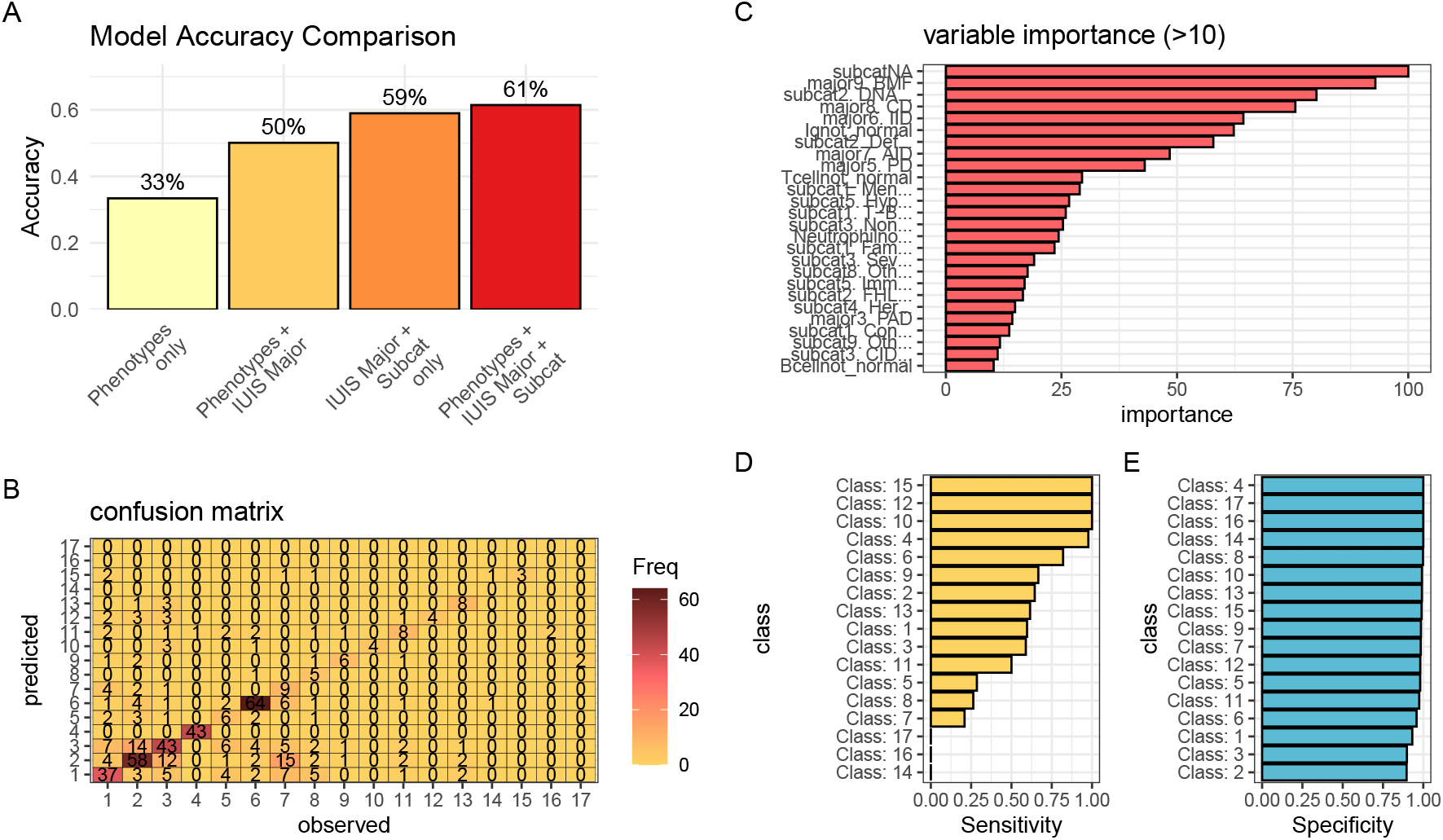
Performance comparison of PID classifiers. Classification predicting PPI cluster membership from IUIS major category, subcategory, and immunological features. (A) Overall accuracy for four rpart models used to predict PPI clustering. The combined model achieves 61.4 % accuracy, exceeding all simpler approaches. Nodes were split to minimize Gini impurity, pruned by cost-complexity (cp = 0.001), and validated via 5-fold cross-validation. (B-E) The summary statistics from the top model are detailed.

### 6.5 Observed variant likelihood is independent of AlphaMis-sense pathogenicity

**Figure S18:**
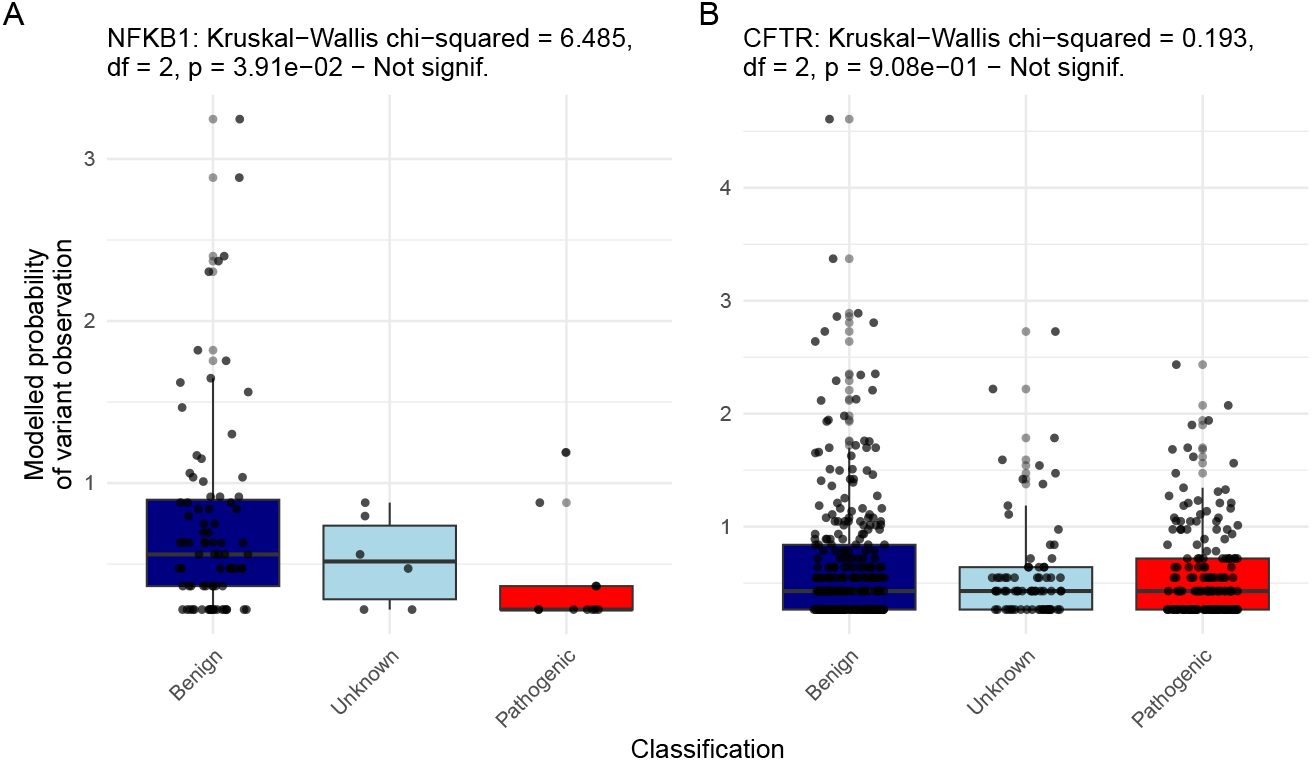
Observed Disease Probability by Clinical Classification with AlphaMissense. The figure displays the Kruskal-Wallis test results for NFKB1 and CFTR, showing no significant differences.

### 6.6 Integration of variant observation probabilities into the IEI genetics framework

**Figure S19:**
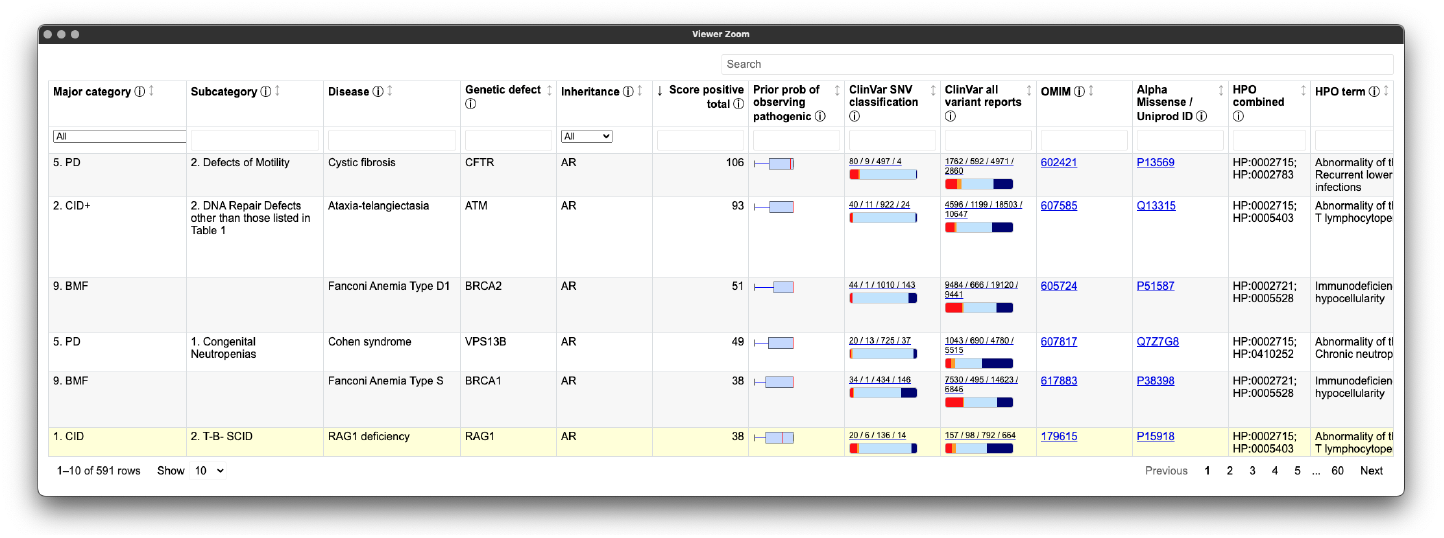
Integration of variant probabilities into the IEI genetics framework. The interface summarises the condensed variant data, with pre-calculated summary statistics and dynamic links to external databases. This integration enables immediate access to detailed variant classifications and prior probabilities for each gene.

